# Optimizing COVID-19 control with asymptomatic surveillance testing in a university environment

**DOI:** 10.1101/2020.11.12.20230870

**Authors:** Cara E. Brook, Graham R. Northrup, Alexander J. Ehrenberg, the IGI SARS-CoV-2 Testing Consortium, Jennifer A. Doudna, Mike Boots

**Affiliations:** Department of Integrative Biology, University of California, Berkeley; Department of Ecology and Evolution, University of Chicago; Center for Computational Biology, College of Engineering, University of California, Berkeley; Innovative Genomics Institute, University of California, Berkeley; Helen Wills Neuroscience Institute, University of California, Berkeley; Memory and Aging Center, Weill Institute for Neurosciences, University of California, San Francisco; Department of Molecular and Cell Biology, University of California, Berkeley; College of Chemistry, University of California, Berkeley; J. David Gladstone Institutes, San Francisco, CA; Howard Hughes Medical Institute, University of California, Berkeley; Department of Biosciences, University of Exeter, Penryn, UK

**Keywords:** COVID-19, asymptomatic surveillance testing, branching process model, university control

## Abstract

The high proportion of transmission events derived from asymptomatic or presymptomatic infections make SARS-CoV-2, the causative agent in COVID-19, difficult to control through the traditional non-pharmaceutical interventions (NPIs) of symptom-based isolation and contact tracing. As a consequence, many US universities developed asymptomatic surveillance testing labs, to augment NPIs and control outbreaks on campus throughout the 2020-2021 academic year (AY); several of those labs continue to support asymptomatic surveillance efforts on campus in AY2021-2022. At the height of the pandemic, we built a stochastic branching process model of COVID-19 dynamics at UC Berkeley to advise optimal control strategies in a university environment. Our model combines behavioral interventions in the form of group size limits to deter superspreading, symptom-based isolation, and contact tracing, with asymptomatic surveillance testing. We found that behavioral interventions offer a cost-effective means of epidemic control: group size limits of six or fewer greatly reduce superspreading, and rapid isolation of symptomatic infections can halt rising epidemics, depending on the frequency of asymptomatic transmission in the population. Surveillance testing can overcome uncertainty surrounding asymptomatic infections, with the most effective approaches prioritizing frequent testing with rapid turnaround time to isolation over test sensitivity. Importantly, contact tracing amplifies population-level impacts of all infection isolations, making even delayed interventions effective. Combination of behavior-based NPIs and asymptomatic surveillance also reduces variation in daily case counts to produce more predictable epidemics. Furthermore, targeted, intensive testing of a minority of high transmission risk individuals can effectively control the COVID-19 epidemic for the surrounding population. Even in some highly vaccinated university settings in AY2021-2022, asymptomatic surveillance testing offers an effective means of identifying breakthrough infections, halting onward transmission, and reducing total caseload. We offer this blueprint and easy-to-implement modeling tool to other academic or professional communities navigating optimal return-to-work strategies.

## Introduction

Non-pharmaceutical interventions (NPIs) to control the spread of infectious diseases vary in efficacy depending on the natural history of pathogen that is targeted [1]. Highly transmissible pathogens and pathogens for which the majority of onward transmission events take place prior to the onset of symptoms are notoriously difficult to control with standard public health approaches, such as isolation of symptomatic individuals and contact tracing [1]. SARS-CoV-2, the causative agent in COVID-19, is a clear example of one of these difficult-to-control pathogens [2]. While the first SARS-CoV was effectively contained via the isolation of symptomatic individuals following emergence in 2002 [3], at the time of this article’s revision, SARS-CoV-2 remains an ongoing public health menace that has infected more than 240 million people worldwide [4]. Though the two coronaviruses are epidemiologically comparable in their original basic reproduction numbers (R_0_) [3], SARS-CoV-2 has evaded control efforts largely because the majority of virus transmission events occur prior to the onset of clinical symptoms in infected persons [2]—in stark contrast to infections with the first SARS-CoV [3]. Indeed, in many cases, SARS-CoV-2-infected individuals never experience symptoms at all [5–8] but, nonetheless, remain capable of transmitting the infection to others [9–13]. Due to the challenges associated with asymptomatic and presymptomatic transmission [10], surveillance testing of asymptomatic individuals has played an important role in COVID-19 epidemic control [14–16]. Asymptomatic surveillance testing is always valuable for research purposes, but its efficacy as a public health intervention will depend on both the epidemiology of the focal infection and the characteristics of the testing regime. Here, we explore the effects of both behavior-based NPIs and asymptomatic surveillance testing on COVID-19 control in a university environment.

In year two of the COVID-19 pandemic, the United States still leads the globe with over 46 million reported cases of COVID-19 [4], and universities across the nation continue to struggle to control epidemics in their campus communities [17]. To combat this challenge in AY2020-2021, colleges adopted a variety of largely independent COVID-19 control tactics, ranging from entirely virtual formats to a mix of in-person and remote learning, paired with strict behavioral regulations, and—in some cases—in-house asymptomatic surveillance testing [18]. In AY2021-2022, asymptomatic surveillance testing continues to play a key role in expanded plans for university reopening [18,19], even on some campuses which also mandate vaccination [20]. In March 2020, shortly after the World Health Organization declared COVID-19 to be a global pandemic [21], the University of California, Berkeley, launched its own pop-up SARS-CoV-2 testing lab in the Innovative Genomics Institute (IGI) [22] with the aim of providing COVID diagnostic services to the UC Berkeley community and underserved populations in the surrounding East Bay region. Though the IGI RT-qPCR-based pipeline was initially developed to service clinical, symptomatic nasopharyngeal and oropharyngeal swab samples [22], the IGI subsequently inaugurated an asymptomatic surveillance testing program for the UC Berkeley community [23], through which—at the time of this revision—over 60,000 faculty, students, and staff in the UC Berkeley community have since been serviced with over 440,000 tests and counting [24]. From June 2020-May 2021, weekly asymptomatic surveillance testing was mandatory for any UC Berkeley community member working on campus; testing requirements were relaxed in May 2021 for those providing proof of vaccination.

Here we developed a stochastic, agent-based branching process model of COVID-19 spread in a university environment to advise UC Berkeley on best-practice approaches for asymptomatic surveillance testing in our community and to offer guidelines for optimal control in university settings more broadly. Previous modeling efforts have used similar approaches to advocate for more frequent testing with more rapid turnaround times at the expense of heightened test sensitivity [14,15] or to weigh the cost-effectiveness of various testing regimes against symptom-based screening in closed university or professional environments [16]. Our model is unique in combining both behavioral interventions with optimal testing design in a real-world setting, offering important insights into efficient mechanisms of epidemic control and an effective tool to optimize control strategies.

## Materials and methods

Our model takes the form of a stochastic branching process model, in which a subset population of exposed individuals (0.5%, derived from the mean percentage of positive tests in our UC Berkeley community [24]) is introduced into a hypothetical 20,000 person community that approximates our university campus utilization goals from spring 2021. With each timestep, the disease parameters for each infected case are drawn stochastically from distributions representing the natural history of the SARS-CoV-2 virus, paired with realistic estimates of the timeline of corresponding public health interventions [2,16,25] (Fig. 1). Our flexible model (Text S1; published here with open-access R-code [26]) allows for the introduction of NPIs for COVID-19 control in four different forms: (1) group size limits, (2) symptom-based isolations, (3) asymptomatic surveillance testing isolations, and (4) contact tracing isolations that follow after cases are identified through screening from symptomatic or asymptomatic surveillance testing (Table 1). Because we focused our efforts on optimal asymptomatic surveillance testing regimes, we did not explicitly model other NPIs, such as social distancing and mask wearing; however, the effects of these behaviors were captured in our representation of R-effective (hereafter, R_E_) for both within-campus and out-of-campus transmission. Since vaccination against SARS-CoV-2 became widely available during the review process of our article (including a vaccine mandate across the University of California school system [27]), we updated our original model to allow for flexible starting conditions that include a variable proportion of vaccinated individuals in a specific university setting. We allowed a randomly selected 5% of vaccinated individuals to become infected and infectious as “breakthrough cases” (consistent with published estimates of vaccine efficacy for the Pfizer-BioNTech mRNA vaccine with the most widespread uptake in the US [28]). For simplicity, we assumed that all infectious individuals were equally transmissible, regardless of vaccination status (though see ‘Discussion’ for future research objectives). After experiencing infection, we further assumed that all individuals became recovered and immune for the remaining duration of our simulations, as our focal timescale of interest (the academic semester) is shorter than most projections of the duration of immunity to SARS-CoV-2 [29,30].

**Fig. 1:**
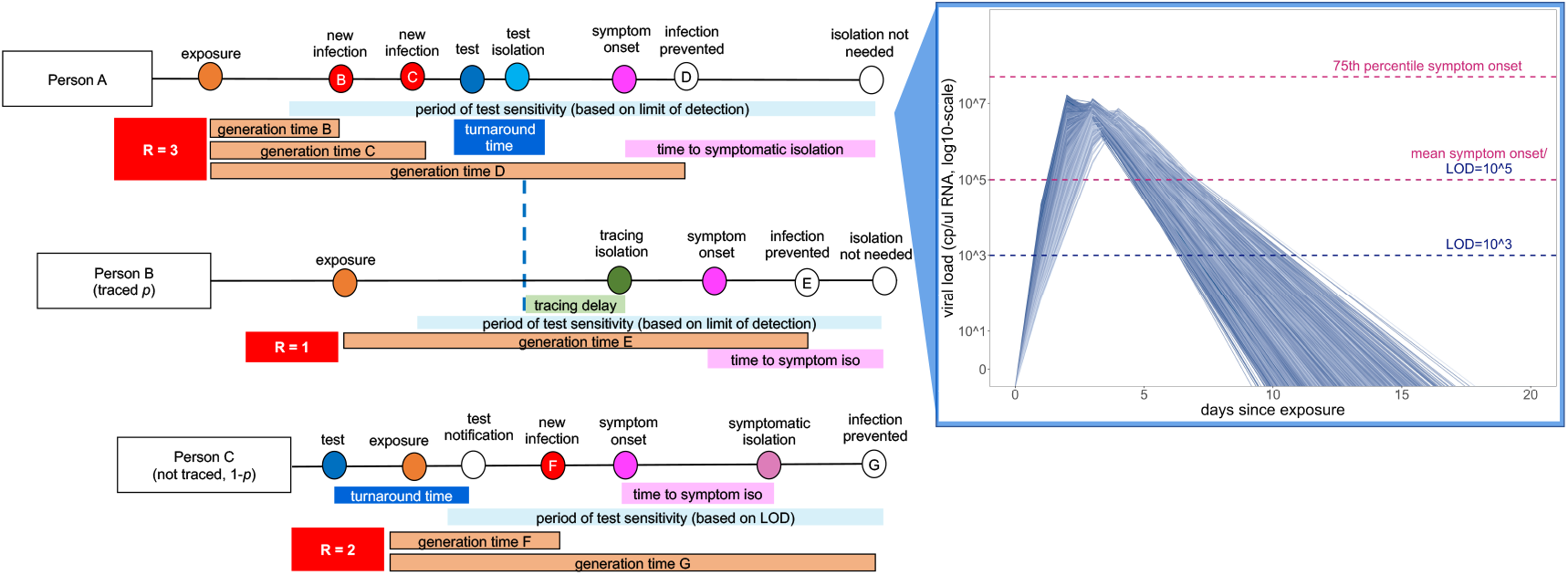
Conceptual schematic of branching process model of SARS-CoV-2 dynamics. Person A is isolated through testing after exposing Person B and Person C. Person B is then isolated through contact tracing, while Person C is not traced but is nonetheless ultimately isolated through symptomatic surveillance. A viral titer trajectory (right) is derived from a within-host viral kinetics model (Text S2)—independent trajectories from 20,000 randomly-selected individuals are shown here to highlight the range of possible variation. The 25^th^ and 75^th^ titer threshold percentile for the onset of symptoms are depicted in pink, such that 32% of individuals modeled in our simulations did not present symptoms. Schematic is adapted in concept from Hellewell et al. (2020) [31].

**Table 1:**
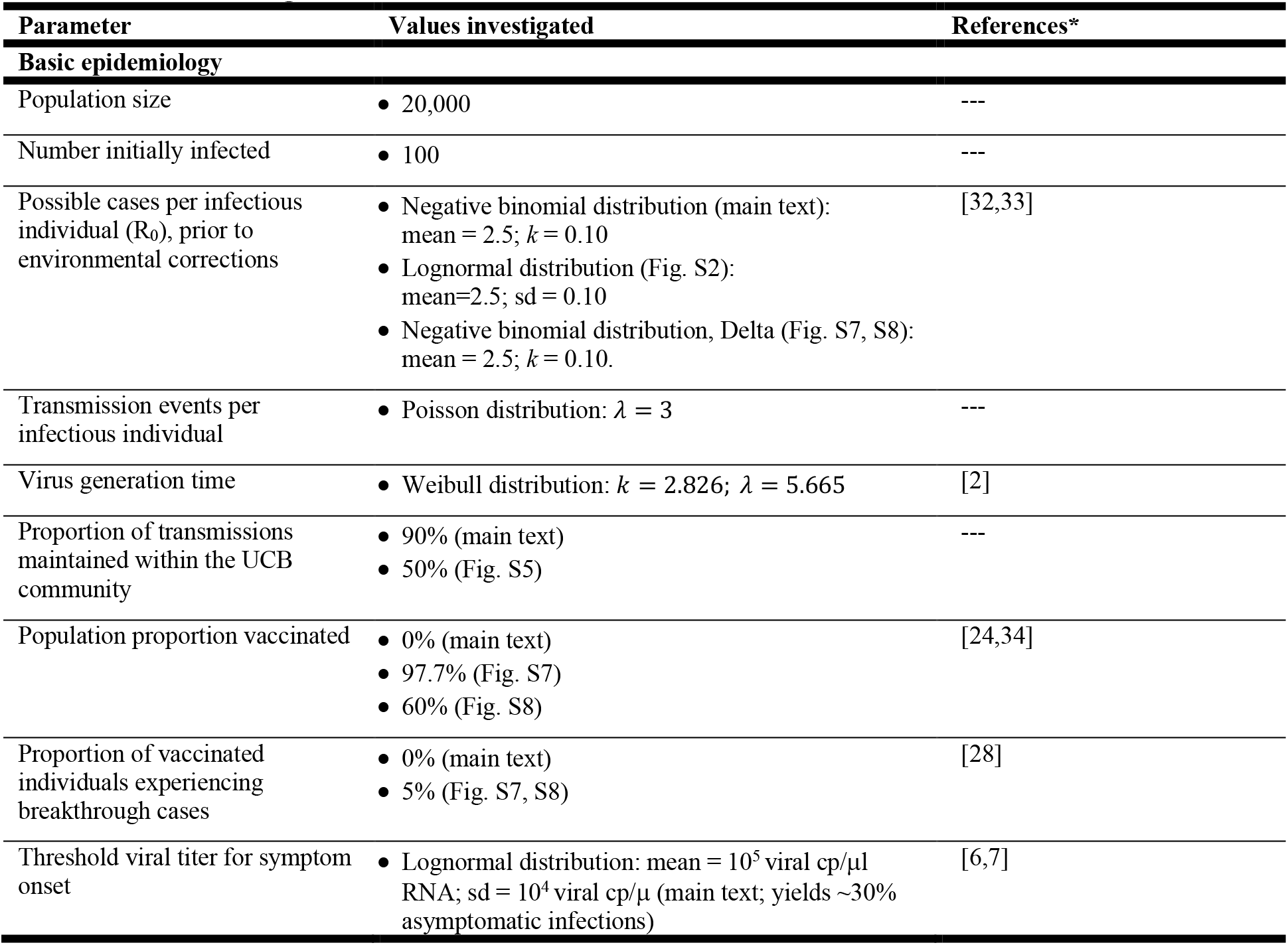
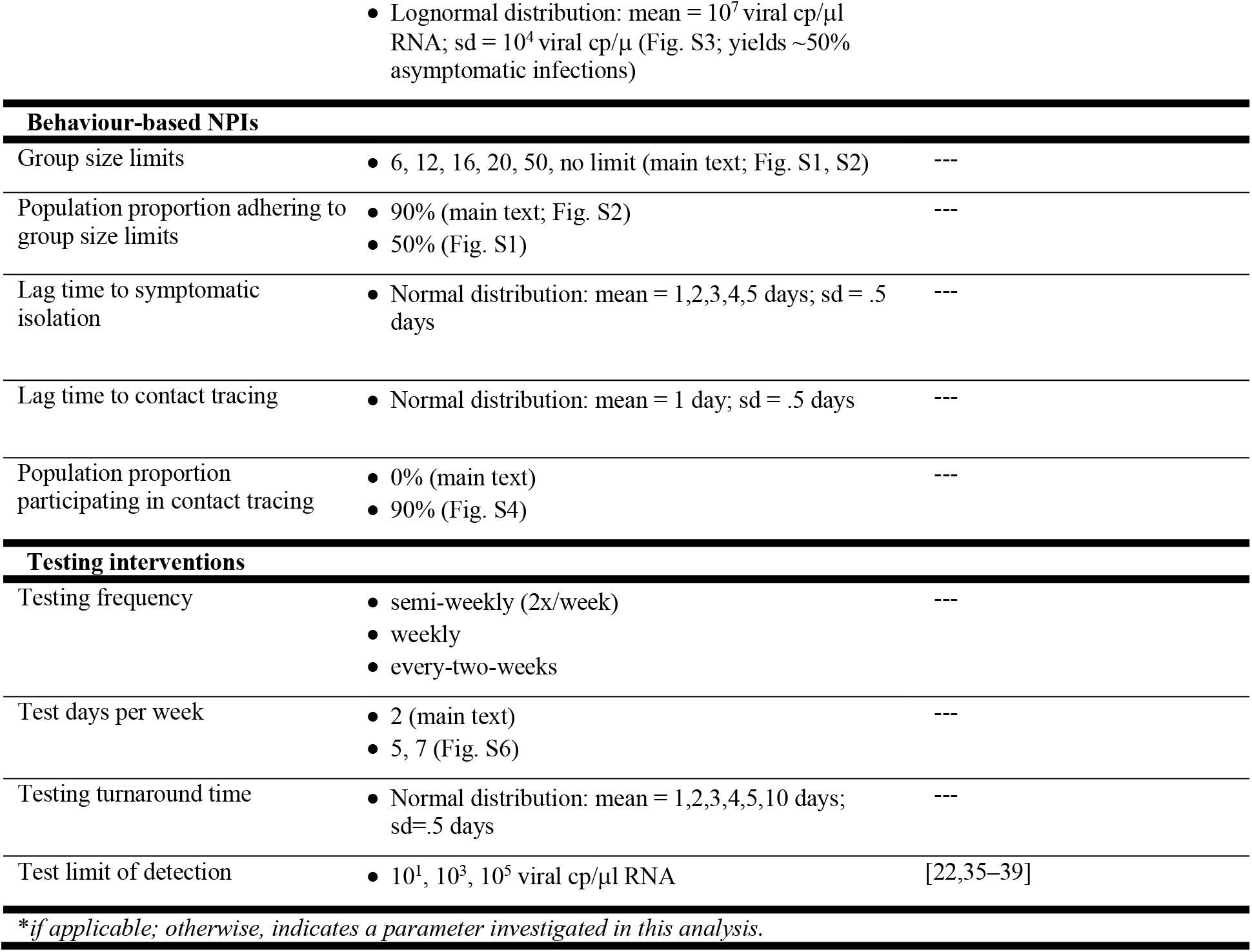
Parameter ranges and interventions included in model.

R_E_ is the product of the pathogen basic reproduction number (R_0_) and the proportion of the population that is susceptible to disease. R_E_ is thus a dynamic value which corresponds to the number of new infections caused by a single infection at a given timepoint within a specified community. We computed an independent R_E_ for each infectious person in our population as a combined result of both heterogeneity in individual infectiousness and heterogeneity in individual contact events that could result in transmission. To determine R_E_, we first drew a value of potential cases for each infectious individual from the SARS-CoV-2 negative binomial distribution for R_0_, estimated to have a mean value of 2.5 and a dispersion parameter (*k*) of 0.10 [32]; in later analyses incorporating highly vaccinated university settings reflective of the reality of AY2021-2022, we shifted the mean to a value of 6 to better approximate the dynamics of highly transmissible variants of concern (e.g. the Delta variant) [33]. Though representation of R_E_ in log-normal vs. negative binomial form will not change the average number of cases generated per epidemic, the negative binomial distribution replicates the dynamics of superspreading events, which are known to play an important role in SARS-CoV-2 dynamics [40–45]. Indeed, there is strong direct empirical evidence that COVID-19 epidemiology exhibits a negative binomial R_E_ across multiple systems [44,46–48]; as few as 10% of infectious individuals may be responsible for 80% of onward SARS-CoV-2 transmissions [49].

After drawing potential cases for each infectious individual, we next hypothesized that most university students would interact predominantly with other students vs. people from the surrounding community and, thus, modeled only a minority (10%) of possible onward transmissions as lost to the external community (e.g. an infectious UC Berkeley community member infects someone outside the UC Berkeley community), though see ‘Results’ for discussion of sensitivity analysis of this assumption.

Next, we assumed that social distancing, masking, and behavioral modifications in our community would modulate dynamics such that some of the remaining 90% (or 50% in sensitivity analyses) of the original R_0_-derived potential infections do not take place. Because we were specifically interested in advising UC Berkeley on group size limits for gatherings, we then drew a number of possible onward transmission events for each infectious individual from a simple Poisson distribution with *λ* = 3, signifying the average number of possible encounters (i.e. cross-household dining, shared car rides, indoor meetings, etc.) per person that could result in transmission. We then use published estimates of the generation time of onward transmission events for SARS-CoV-2 infection [2] to draw event times for these encounters and distributed each infectious person’s original number of R_0_-derived potential cases among these events at random. This ensured that multiple transmissions were possible at a single event; the most extreme superspreading events occur when persons with heterogeneously high infectiousness draw a large number of potential cases, which are concentrated within a relatively small number of discrete transmission events. When we imposed group size limit NPIs in our model, we truncated case numbers for each event at the intervention limit.

For each infectious individual, we additionally generated an independent virus trajectory, using a within-host viral kinetics model for SARS-CoV-2 upper respiratory tract infections, structured after the classic target cell model [50–53] (Text S2). From each independent virus trajectory, we inferred a time-varying transmissibility, modeled as a Michaelis-Menten-like function of viral load [53]. We fixed the within-host viral kinetics model constant, *θ*, at a value that allowed for a ∼50% probability of infection occurring per transmissible contact event at an infectious individual’s peak viral load [53]. Because all possible onward transmissions were assigned an event generation time, we next evaluated the viral load of the infectious person at the time of each potential transmission to determine whether or not it actually occurred. By these metrics, our original R_0_-derived possible cases were halved, such that R_E_, the number of average onward infections caused by a single infectious person in the UC Berkeley community, was reduced to just over one (R_E_=1.05), or just under three (R_E_=2.94) in the case of Delta variant simulations, consistent with published estimates of Bay Area R_E_ and initial asymptomatic test results in our community from the first year of the pandemic [24,54]. The majority of modelled transmission events occurred when the infectious host had higher viral titers, thus biasing new case generations towards earlier timesteps in an individual’s infection trajectory, often occurring prior to the onset of symptoms as is realistic for COVID-19 [25] (Fig. 1).

In addition to modulating the probability of onward transmission events, each infectious individual’s virus trajectory additionally allowed us to compute a timing of symptom onset, which corresponded to the timepoint at which an individual’s virus trajectory crossed some threshold value for presentation of symptoms. We drew each threshold randomly from a log-normal distribution with a mean of 10^5^ virus copies per μl of RNA; by these metrics, roughly 32% of our modeled population presented as asymptomatic, in keeping with published estimates for SARS-CoV-2 [6,7]. Using each infectious individual’s viral load trajectory, we were next able to compute a period of test sensitivity, corresponding to the time during which viral load is high enough for detection by the virus test in question, based on the modeled limit of detection. Asymptomatic surveillance testing results in higher “false-negative” test results both very early and very late in infection when viral loads are below the detection limit for the adopted assay [55] (Fig. 1), though most tests should reliably detect infectious cases with viral titers >10^6^ cp/μl [56–58]. We explored dynamics across a range of published values for test limits of detection: 10^1^, 10^3^, and 10^5^ virus copies per μl of RNA. The IGI’s RT-qPCR-based testing pipeline has a published sensitivity of 1 cp/μl [22], while the majority of SARS-CoV-2 RT-qPCR tests nationally are reliable above a 10^3^ cp/μl threshold [35]; less-sensitive antigen-based and LAMP assays report detection limits around 10^5^ cp/μl [36,37]. Some commercially-available COVID-19 test kits detection limits in TCID_50_/ml, which corresponds to the median tissue culture infectious dose, roughly approximating a threshold for the infectious viral load. Though exact values will vary depending on the virus, cell type, and assay conditions, a 100 TCID_50_/ml limit of detection for SARS-CoV-2 has been shown to correspond to a viral load detection limit between 10^2^ and 10^3^ cp/μl RNA [38,39]. For reference, the Abbot BinaxNOW™ COVID-19 Ag card reports a limit of detection of 140.6 TCID_50_/ml (between 10^2^ and 10^3^ cp/μl RNA), while the QuickVue At-Home COVID-19 test reports a limit of detection of 1.91×10^4^ TCID_50_/ml (between 10^4^ and 10^5^ cp/μl RNA).

In addition to within-community transmissions, all individuals in the modeled population were also subjected to a daily hazard (0.25% in standard model runs and 0.60% in Delta variant runs) of becoming infected from an external source, based on published estimates of R_E_ and COVID-19 prevalence in Alameda County [54,59]. We report the mean results of 100 stochastic runs of each proposed intervention.

## Results

### Comparing behavioral NPIs for COVID-19 control

We first ran a series of epidemic simulations using a completely mixed population of 20,000 individuals subject to the infection dynamics outlined above to compare and contrast the impacts of our four NPIs on COVID-19 control. We introduced an initial population of 100 infectious individuals (0.5%) at timestep 0 and compared the effects of a single intervention on epidemic trajectories after the first 50 days of simulation. Less intensive or intervention-absent scenarios allowed infectious cases to grow at unimpeded exponential rates, rapidly exhausting our susceptible supply and making it necessary to compare results at a consistent (and early) timepoint in our simulated epidemics.

As a consequence of our representation of R_E_ in negative binomial form, we first considered the COVID-19 control effectiveness of group size limits on in-person gatherings, which doubled as upper thresholds in transmission capacity (Fig. 2). Assuming that 90% of the modeled population adhered to assumed group size regulations, we found that limiting outdoor gatherings to groups of six or fewer individuals saved a mean of ∼7,900 cases per 50-day simulation (in a 20,000 person population) and corresponded to an R_E_ reduction of nearly 0.20 (reducing R_E_ from 1.05 to subclinical 0.86; Fig. 2; Dataset S1). By contrast, a large group size limit of 50 persons had almost no effect on epidemic dynamics; under published estimates of SARS-CoV-2 negative binomial R_E_ [32], a group size limit of 50 will restrict transmission from only 0.00039% of infectious individuals (Fig. 2). Intriguingly, in sensitivity analyses exploring assumptions of only 50% adherence to group size limits, we witnessed larger caseloads only at group size limits of 16 or fewer individuals (Fig. S1); at group sizes of 20 or more individuals, density limits were so ineffective already that reducing adherence had no power to further undermine the intervention’s impacts. Gains in epidemic control from group size limits resulted from avoidance of superspreading events, an approach that was effective for negative binomial but not log-normal representations of R_E_ that lack the transmission “tail” characteristic of a superspreader distribution [45] (Fig. S2). Importantly, by avoiding superspreading events, group size limits also reduced variance in daily case counts, yielding more predictable epidemics, which are easier to control through testing and contact tracing [2,25,31]. Over the July 4, 2020 weekend, asymptomatic surveillance testing resources in our UC Berkeley community were overwhelmed and containment efforts challenged after a single superspreading event on campus [60].

**Fig. 2:**
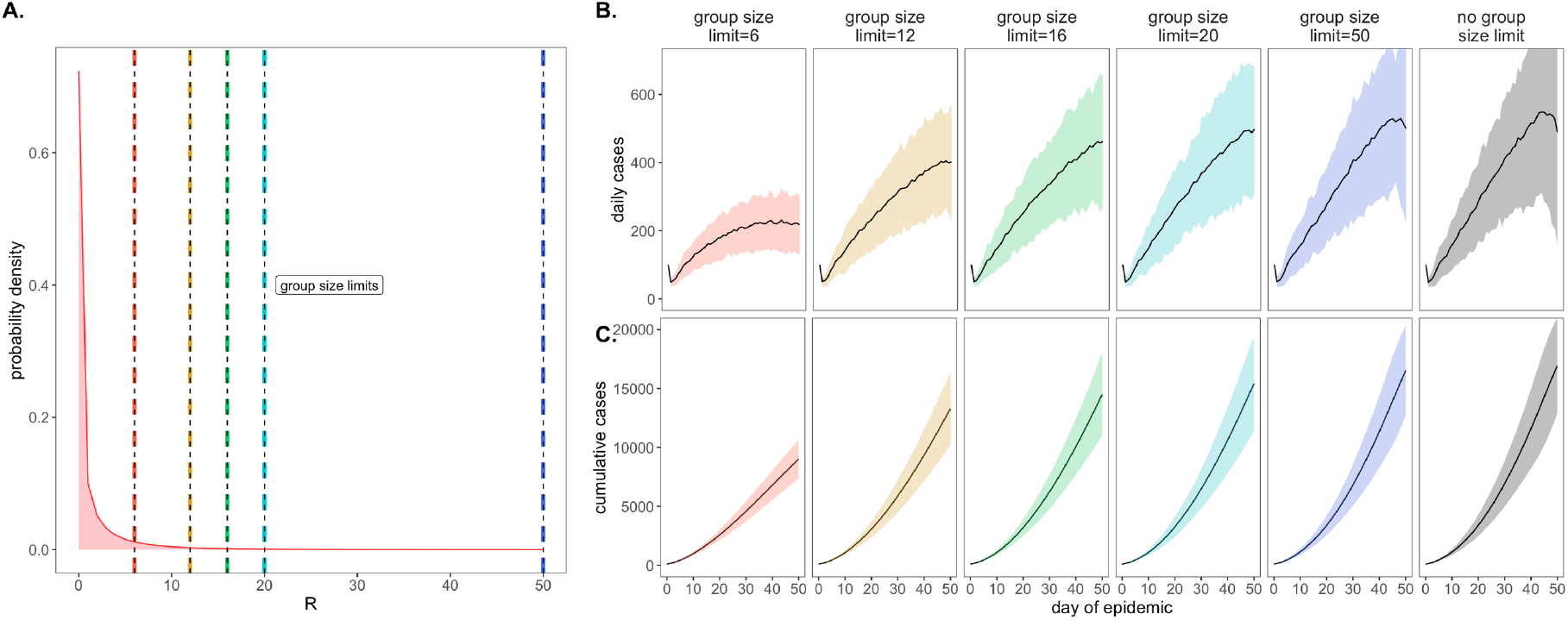
Effects of group size limits on COVID-19 dynamics. **A**. Negative binomial R_E_ distribution with mean = 1.05 and dispersion parameter (k) = 0.10. The colored vertical dashes indicate group size limits that ‘chop the tail’ on the R_E_ distribution; for 90% of the population, coincident cases allocated to the same transmission event were truncated at the corresponding threshold for each intervention. **B**. Daily new cases and, **C**. Cumulative cases, across a 50-day time series with 95% confidence intervals by standard error depicted under corresponding, color-coded group size limits.

We next investigated the impacts of variation in lag time to self-isolation post-symptom onset for the just under 70% of individuals likely to present with COVID-19 symptoms in our modeled population (Fig. 3). At UC Berkeley, all essential students, faculty, and staff must complete a digital ‘Daily Symptom Screener’ before being cleared to work on campus; here, we effectively modeled the delay post-initial symptom onset to the time at which each individual recognizes symptoms sufficiently to report to the Screener and isolate. For each infected individual in our population, we drew a symptom-based isolation lag from a log-normal distribution centered on a mean of one to five days, assuming the entire population to be compliant with the selected lag.

**Fig. 3:**
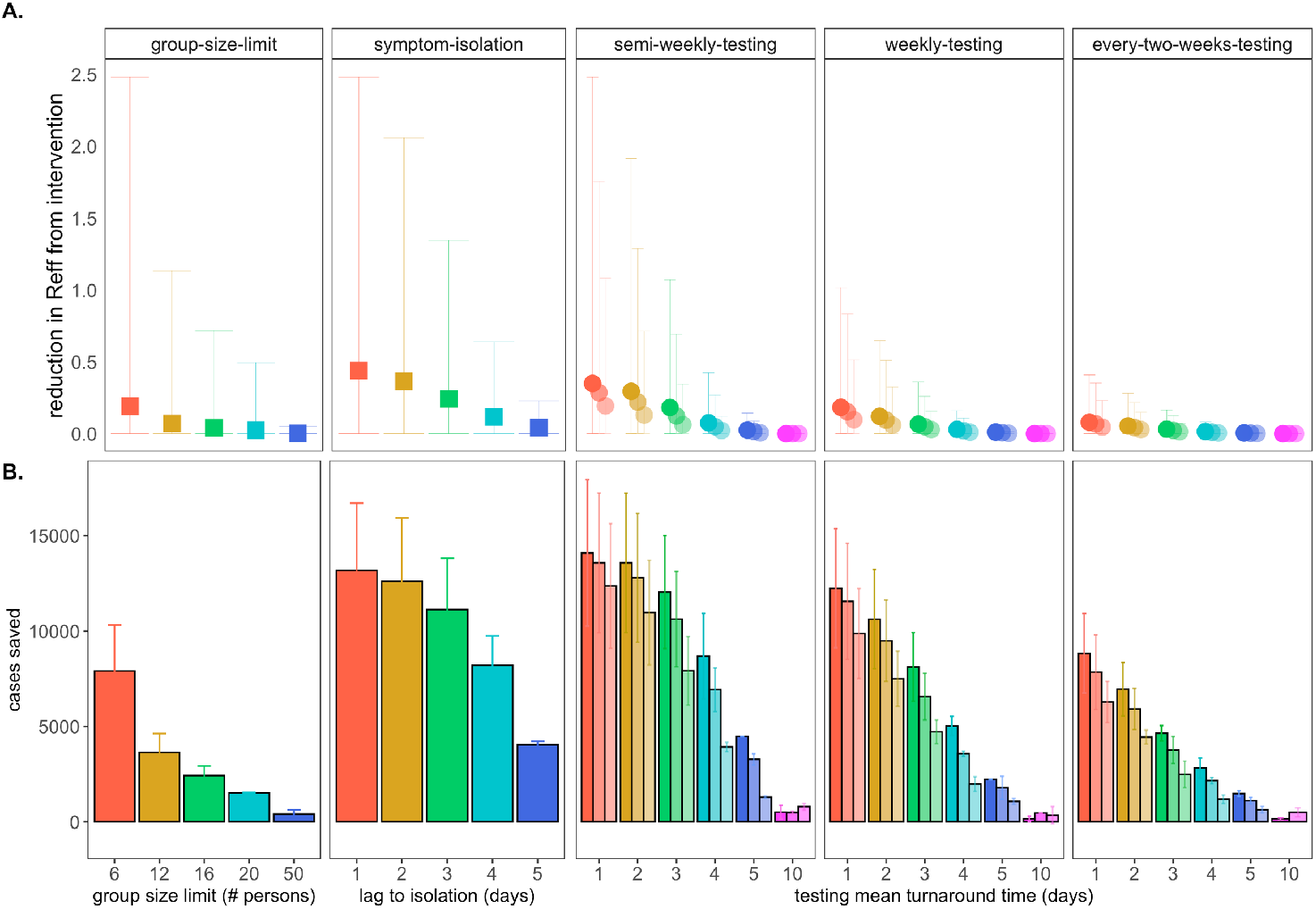
Impacts of NPIs on COVID-19 control. **A**. Mean reduction in R_E_* and **B**. cumulative cases saved across 50-day simulated epidemics under assumptions of differing non-pharmacological interventions (NPIs). NPIs are color-coded by threshold number of persons for group-size limits, lag-time for symptom-based isolations, and mean turnaround time from test positivity to isolation of infectious individuals for testing isolations. For testing isolations, shading hue corresponds to test limit of detection with the darkest colors indicating the most sensitive tests with a limit of detection of 10^1^ virus copies/μl of RNA. Progressively lighter shading corresponds to limits of detection = 10^3^, 10^5^, and 10^7^ cp/μl. *Note: R_E_ reduction (panel A) is calculated as the difference in mean R_E_ in the absence vs. presence of a given NPI. The upper confidence limit (uci) in R_E_ reduction is calculated as the difference in uci R_E_ in the absence vs. presence of NPI. In our model, mean R_E_ in the absence of NPI equals 1.05 and uci R_E_ in the absence of NPI equals 8.6.

By these metrics, a rapid, one day lag in symptom-based isolation was the fourth-most effective intervention in our study, with a mean of more than 13,100 cases saved in a 50-day simulation (again, in a 20,000 person population), corresponding to an R_E_ reduction of 0.67, from 1 to 0.38 (Dataset S1). Longer lag times to isolation produced less dramatic results, but even an average five-day lag to isolation post-symptom onset nonetheless yielded more than 4,000 cases saved and reduced R_E_ by a mean of 0.06. The efficacy of symptom-based isolation decreased at higher virus titer thresholds for symptom onset, corresponding to a higher asymptomatic proportion (∼50%) of the population (Fig. S3); some empirical findings suggest that these higher titer thresholds for symptom onset may more accurately reflect COVID-19 epidemiology [61]. Because both group size limits and daily screening surveys to facilitate symptom-based isolation can be implemented without expending substantial resources, we advocate for these two approaches as particularly cost-effective COVID-19 control strategies for all university and small community environments—especially those lacking an on-site asymptomatic surveillance testing lab.

### Comparing asymptomatic surveillance testing for COVID-19 control

Our primary motivation in developing this model was to advise UC Berkeley on best-practices for asymptomatic surveillance testing. As such, we focused efforts on determining the most effective use of testing resources by comparing asymptomatic surveillance testing across a range of approaches that varied test frequency, test turnaround time (the time from which the test was administered to the timing of positive case isolation), and test sensitivity (based on the limit of detection).

We compared all permutations of asymptomatic surveillance testing, varying test frequency across semi-weekly, weekly, and every-two-week regimes, investigating turnaround time across delays of one to five and ten days, and exploring limits of detection of 10^1^, 10^3^, and 10^5^ virus copies per μl of RNA. These test frequency regimes reflect those considered by UC Berkeley administrators throughout the pandemic: from August-December 2020 and January-April 2021, UC Berkeley undergraduates residing in university residence halls were subject to compulsory semi-weekly asymptomatic surveillance testing, while all other campus community members were permitted to take part in voluntary testing with a recommended weekly or every-two-week frequency. After vaccines became widespread (and eventually mandated), testing requirements for vaccinated undergraduates in residence halls were reduced to once a month. Turnaround time values in our model reflect the reality in range of testing turnaround times from in-house university labs like that at UC Berkeley to institutions forced to outsource testing to commercial suppliers [62], and limits of detection span the range in sensitivity of available SARS-CoV-2 tests [22,35–37].

Across testing regimes broadly, we found test frequency, followed by turnaround time, to be the most effective NPIs, with limit of detection exerting substantially less influence on epidemic dynamics, consistent with findings published elsewhere [14,15]. The top three most effective NPIs in our study corresponded to semi-weekly testing regimes with one- and two-day turnaround times across 10^1^ and 10^3^ cp/μl limits of detection. These three scenarios yielded mean cases saved ranging from just over 14,000 to just over 13,500 in the first 50 days of simulation and produced an R_E_ reduction capacity between 0.97 and 0.80 (Fig. 3; Dataset S1). Halving test frequency to a weekly regimen, under assumptions of turnaround time=1 day and limit of detection=10^1^, resulted in a nearly 48% decrease in the NPI’s R_E_ reduction capacity. By comparison, a single extra day lag from one to two-day turnaround time under semi-weekly testing conditions at limit of detection=10^1^ cp/μl yielded a modest 16% decrease in R_E_ reduction capacity. However, longer delays in turnaround time of up to ten days or more—not unusual in the early stages of the COVID-19 pandemic [62]—were not significantly different from scenarios in which no intervention was applied at all. This outcome results from the rapid generation time of SARS-CoV-2 [2]; most infectious individuals will have already completed the majority of subsequent transmissions by the time a testing isolation with a 10-day turnaround time is implemented. Nonetheless, encouragingly, reducing test sensitivity from 10^1^ to 10^3^ under a semi-weekly, turnaround time=1 day regime decreased R_E_ reduction capacity by only 18%, offering support to advocates for more frequent but less sensitive tests [63] but also highlighting the added benefit incurred when university testing labs, like that at UC Berkeley, are able to provide both frequent and sensitive PCR-based testing.

Addition of a contact tracing intervention, in which 90% of infectious contacts were traced and isolated within a day of the source host isolation, to NPI scenarios already featuring either symptom-based or asymptomatic surveillance testing isolation enhanced each intervention’s capacity for epidemic control (Fig. S4). Of note, contact tracing boosted performance of some of the poorest performing testing interventions, such that even those previously ineffective asymptomatic surveillance regimens with 10-day turnaround time nonetheless averted cases and significantly reduced R_E_ when infectious contacts could be isolated. For a semi-weekly testing regime at limit of detection =10^1^ cp/μl and turnaround time=10 days, the addition of contact tracing increased mean cases saved from ∼510 to >8,600 and increased R_E_ reduction capacity from 0.000080 to 0.27 (Dataset S2).

### Optimizing combined NPIs for COVID-19 control

Our modeled simulations suggested that it is possible to achieve largely equivalent gains in COVID-19 control from NPIs in the form of group size limits, symptom-based isolations, and asymptomatic surveillance testing isolations—though gains from symptom-based behavioral isolations were jeopardized under assumptions of a higher proportion of asymptomatic individuals (Fig. S3). Nonetheless, the most effective interventions were realized when behavioral control mechanisms were *combined* with asymptomatic surveillance testing (Fig. 4). Assuming a one day turnaround time and 10^1^ cp/μl limit of detection, we found that adding (a) contact tracing with 90% adherence and a one-day lag, plus (b) symptom-based isolation with a one-day lag, plus (c) a group size limit of twelve persons to an every-two-week asymptomatic surveillance testing regimen could elevate the R_E_ reduction capacity from 0.22 to 0.83 and almost double the ∼6,600 cases saved from the testing intervention alone (Dataset S3). Combining interventions enabled less rigorous testing regimes to rival the effectiveness of semi-weekly asymptomatic surveillance testing without expending additional resources. In addition, combining interventions resulted in less variation in the cumulative case count, as many layers of opportunity for infection isolation helped limit the likelihood of a superspreading event spiraling out of control. Sensitivity analyses indicated that our findings were largely robust to assumptions of exacerbated insularity in university settings (e.g. when only 1% of transmissions were lost to the outside) but that the impacts of combined interventions were reduced under sensitivity analyses exploring a higher proportion (e.g. 50%) of transmissions lost to the external community (Fig. S5), as interventions can only be applied within the closed campus. These findings highlight the vulnerability of any community public health control measure to disease introductions from beyond the sphere of control. On a macroscale, isolated countries like New Zealand have struggled with this challenge across the course of the COVID-19 pandemic [64].

**Fig. 4:**
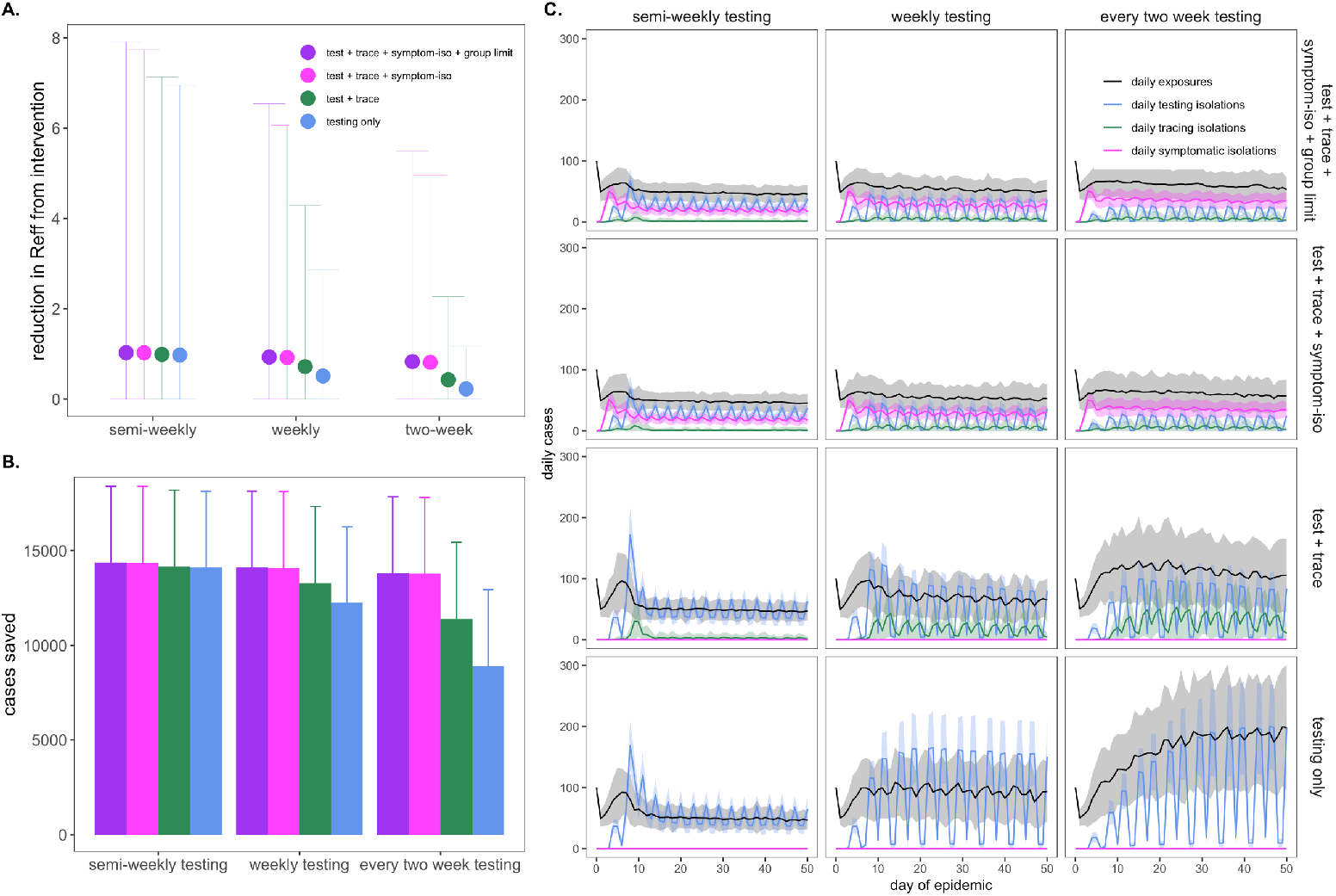
Combining behavioral and asymptomatic surveillance testing NPIs for COVID-19 control. **A**. Mean reduction in R_E_*, **B**. cumulative cases saved, and **C**. daily case counts for the first 50 days of the epidemic, across regimes of differing testing frequency and a combination of asymptomatic surveillance testing, contact tracing, symptomatic isolation, and group size limit interventions. All scenarios depicted here assumed test turnaround time, symptomatic isolation lags, and contact tracing lags drawn from a log-normal distribution with mean=one day. Limit of detection was fixed at 10^1^ and group size limits at 12. Dynamics shown here are from simulations in which testing was limited to two test days per week. *Note: R_E_ reduction (panel A) is calculated as the difference in mean R_E_ in the absence vs. presence of a given NPI. The upper confidence limit (uci) in R_E_ reduction is calculated as the difference in uci R_E_ in the absence vs. presence of NPI. In our model, mean R_E_ in the absence of NPI equals 1.05 and uci R_E_ in the absence of NPI equals 8.6.

Finally, we also experimented with varying the distribution of days allocated to asymptomatic surveillance testing, without changing the frequency with which each individual was tested. Specifically, we explored semi-weekly, weekly, and every-two-week testing regimens in which tests were administered across two, five, and seven available testing days per week. More broadly distributed test days corresponded to fewer tests per day at a population level but, as with more intervention layers, resulted in less variation in the cumulative total cases because testing isolations more closely tracked daily exposures (Fig. S6).

### Modeling COVID-19 dynamics in the campus community

We next sought to advise the IGI on asymptomatic surveillance testing strategies explicitly by simulating epidemics in a more realistic, heterogeneous population modeled after the UC Berkeley campus community in the spring semester of AY2020-2021 (Fig. 5). To this end, we subdivided our 20,000 person university population into a 5,000 person “high transmission risk” cohort and a 15,000 person “low transmission risk” cohort, assuming “high transmission risk” status to correspond to individuals (such as undergraduates), living in high density housing with a majority of contacts (90%) concentrated within the UCB community and “low transmission risk status” to correspond to individuals (such as faculty members or postdoctoral scholars) with only limited contacts (40%) in the UCB community. We imposed a 12-person group size limit (with 90% adherence) on the population as a whole, as recommended by the City of Berkeley Public Health Department in the early months of the pandemic [65], and assumed a one-day average lag in symptom-based isolation for all cohorts. To add additional realism, we enrolled only 50% of each transmission risk group in our modeled asymptomatic surveillance testing program (to mimic adherence—though asymptomatic surveillance testing is compulsory for undergraduates residing in residence halls at UC Berkeley [24]). We assumed that 95% efficacy in contact tracing (with a mean tracing delay of one day) for those enrolled in our asymptomatic surveillance program but only 50% efficacy for those not enrolled; UC Berkeley has encouraged all community members to enroll in the ‘CA Notify’ digital contract tracing app developed by Apple and Google [66]. For all testing interventions, we assumed limit of detection=10^1^ cp/μl and turnaround time=2 days, the average for the IGI asymptomatic surveillance testing lab [22].

**Fig. 5:**
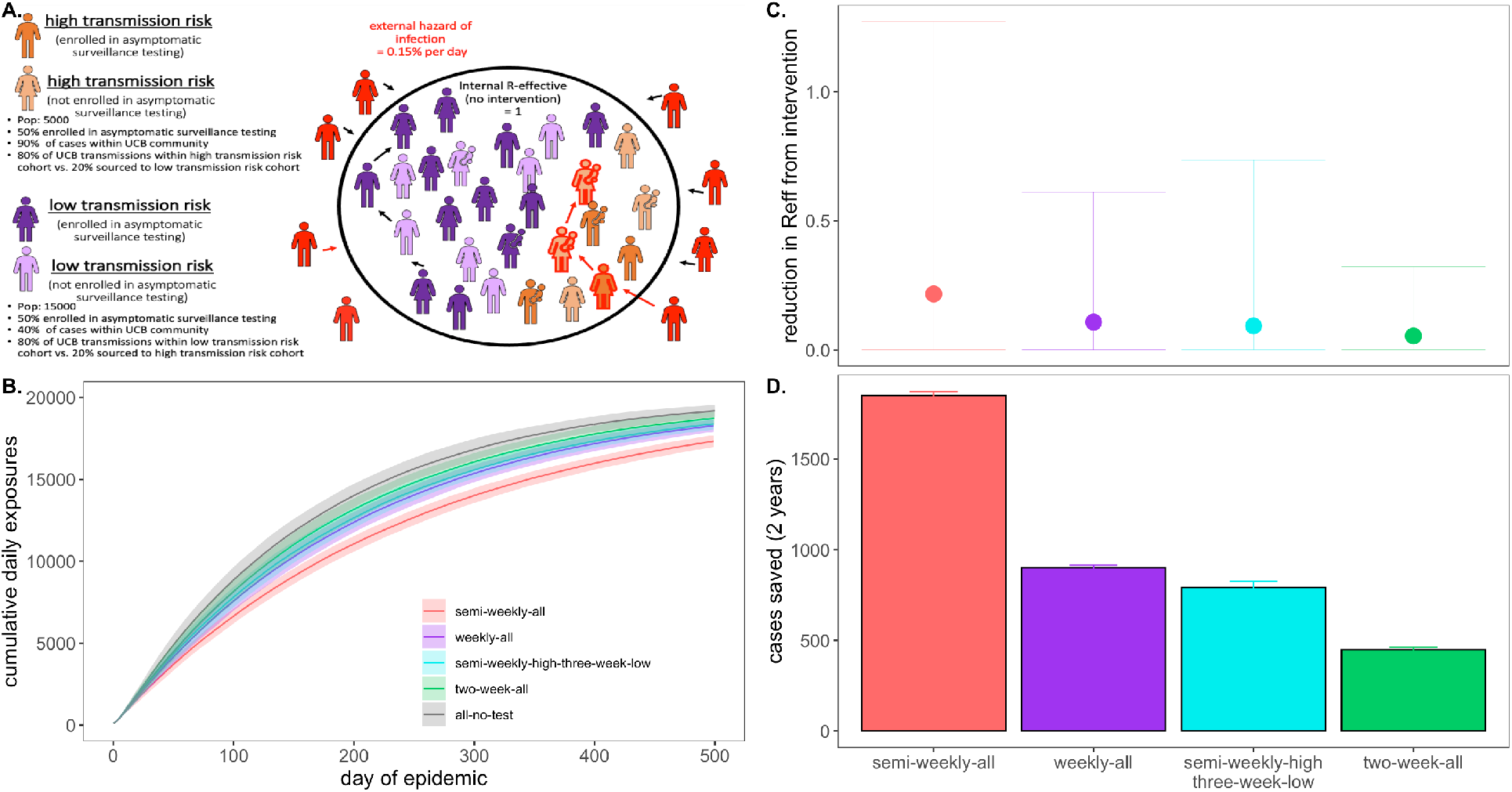
Targeted testing of high transmission risk cohorts in a heterogenous population. **A**. Schematic of transmission risk group cohorts in the heterogenous model. The population is divided into 5,000 “high transmission risk” and 15,000 “low transmission risk” individuals, for which, 90% and 40% of the proportion of transmission events take place within the UC Berkeley community, respectively. Of those transmission events within the Berkeley community, the majority (80%) are restricted within the same transmission risk group as the infector, while 20% are sourced to the opposing risk group. Half of each cohort is assumed to be enrolled in asymptomatic surveillance testing and subjected to the differing test frequency regimes depicted in panels **B**. through **D**. Panel **B**. shows the progression of cumulative cases across 730 days of simulation for each testing regime, while panel **C**. and **D**. give, respectively, the reduction in R_E_* and the total cases saved achieved by each test regime vs. a no intervention baseline. *Note: R_E_ reduction (panel A) is calculated as the difference in mean R_E_ in the absence vs. presence of a given NPI. The upper confidence limit (uci) in R_E_ reduction is calculated as the difference in uci R_E_ in the absence vs. presence of NPI. In our model, mean R_E_ in the absence of NPI equals 1.05 and uci R_E_ in the absence of NPI equals 8.6.

We found that targeted, semi-weekly testing of 50% of individuals in the high transmission risk cohort, paired with every-three-week testing of enrolled individuals in the low transmission risk cohort yielded mean R_E_ reduction and cumulative cases saved on par with that achieved from weekly testing (and better than that achieved from every-two-week testing) of all enrolled individuals in the population at large (Fig. 5). Targeting the highest transmission-risk populations with testing allows practitioners to save valuable resources while simultaneously controlling the epidemic for the entire community. Importantly, while mean R_E_ reduction and cumulative cases were largely comparable between the targeted, semi-weekly testing regiment and the untargeted, weekly regimen, the observed variance in intervention efficacy (Fig. 5C) was substantially greater for the targeted scenario, in which the low transmission risk cohort was only tested once every three weeks. This results from a higher probability that a rare superspreading event could occur in the infrequently monitored low transmission risk cohort, thus reaffirming our previous observation that more frequent asymptomatic surveillance testing regimens result in more predictable—and easier to control—epidemics.

Notably, irrespective of intervention, the diminished transmissibility of the “low transmission risk” population in this heterogeneous model structure greatly reduced epidemic spread in subsequent simulations as compared with those presented previously in the perfectly mixed environment; as a result, we here compared interventions after 500 days of simulation, rather than 50. The heightened realism of our heterogenous population generated slow-moving epidemics more closely resembling those we witnessed in our university environment across AY2020-2021.

### Modeling vaccinated environments

During the time in which this article was under review, COVID-19 vaccines became widely available in the US, and the University of California system issued a vaccine mandate for students and staff across all of its campuses, including UC Berkeley [27]. Simultaneously, the highly transmissible Delta variant (R_0_ ∼ 6 [33]) took hold as the most widespread SARS-CoV-2 lineage in the United States [67]. To address this new reality, we ran additional simulations of our original, single-population, university testing model, comparing the mosaic of possible interventions exhibited in Fig. 4 under assumptions of R_0_ = 6 in university settings in which a variable proportion of the student population was vaccinated. Specifically, we compared simulations in a population that was only 60% vaccinated (reflecting the student population of the University of Alabama, Tuscaloosa, a comparably sized public university to UCB but without a vaccine mandate, at the time of writing [34]) to simulations in a population that was 97.7% vaccinated (reflecting the UC Berkeley undergraduate population at the time of writing [24]). Over 1,000 US universities and colleges have now issued guidelines mandating vaccination (with some exceptions) for on-campus study [68].

In these new simulations, testing, tracing, symptomatic isolation, and group size limit NPIs continued to have scalable impacts on COVID-19 dynamics within each respective university setting (Fig. S7-S8). Baseline R_E_ under Delta variant assumptions in 60% vaccinated populations without behavior- or testing-based interventions was higher than baseline R_E_ in unvaccinated populations under standard transmission assumptions (1.12 vs. 1.05). Nonetheless, behavior- and testing-based NPIs easily controlled epidemics in a less susceptible population (Fig. S7). Averted cases were fewer because fewer infections occurred altogether in the partially-vaccinated population. Daily variance in exposure rate narrowed and differences in impact between interventions of variable intensity were less extreme in this more mild epidemic scenario, a pattern even more more pronounced in simulations assuming a 97.7% vaccinated population. Under assumptions of near-complete vaccination and Delta transmission, baseline R_E_ equaled 0.17, and a testing only intervention with an every-two-week frequency was sufficient to avert the majority of onward transmission in the system (Fig. S8). Our findings offer support for some university policies which continue to mandate asymptomatic surveillance testing even for vaccinated individuals [20], as even modest surveillance efforts still effectively reduced R_E_ and averted cases in highly vaccinated settings. Our model is structured such that future work could investigate the impact of disparate population sizes, distinct R_0_ values reflective of variable contact patterns, and unique vaccination proportions in heterogeneous subgroups within a larger community on longterm epidemic control.

## Discussion

We built a stochastic branching process model of SARS-CoV-2 spread in a university environment to advise UC Berkeley on best-practice strategies for effective asymptomatic surveillance in our pop-up IGI testing lab—and to offer a model for other institutions attempting to control the COVID-19 epidemic in their communities. While previous work has explored the isolated effects of specific NPIs—including group association limits [45], symptomatic isolation [2,14–16,25,31], asymptomatic surveillance testing [14–16], and contact tracing [2,25,31]—on COVID-19 control, ours is unique in investigating these interventions simultaneously in a realistic and easily applicable setting. We offer an easy-to-implement modeling tool that can be applied in other educational and workplace settings to provide NPI recommendations tailored to the COVID-19 epidemiology of a specific environment.

Results from our analysis of behavior-based NPIs support previous work [2,14–16,25,31,45] in showing that stringent group size limitations to minimize superspreading events and rapid symptom-based isolations offer an effective means of epidemic control in the absence of asymptomatic surveillance testing resources. However, because of the unique natural history of the SARS-CoV-2 virus, for which the majority of transmission events result from asymptomatic or presymptomatic infections [2,31], symptom-based NPIs cannot reduce epidemic spread completely, and small community environments will always remain vulnerable to asymptomatic case importation. Moreover, symptom-based NPIs pose less effective means of epidemic control under scenarios assuming a higher proportion of asymptomatic individuals; empirical evidence suggests that SARS-CoV-2 infection may result in asymptomatic infection in up to nearly 70% of the population in select environments [61]. For this reason, our results emphasize the importance of asymptomatic surveillance testing to prevent ongoing epidemics in universities and other small community environments. As more data becomes available on both the proportion of asymptomatic infections and their contributions to SARS-CoV-2 transmission, the relative importance of group size interventions, symptom-based isolation, and asymptomatic surveillance testing in different epidemiological contexts will be possible to determine from our modeling framework.

As with behavioral interventions, our exploration of optimal asymptomatic surveillance testing regimes supports findings that have been published previously but with some key extensions and critical novel insights. As has been recently highlighted [14,15], we find that the most cases are saved under asymptomatic testing regimes that prioritize heightened test frequency and rapid turnaround time over test sensitivity. Importantly, we extend previous work to highlight how more rigorous testing regimes—and those combined with one or more behavioral interventions—greatly reduce variance in daily case counts, leading to more predictable epidemics. We find that the reduction in daily case variation is even more pronounced when test regimes of equivalent frequency are distributed more broadly in time (i.e. tests are offered across more days of the week), thus minimizing the likelihood of compounding transmission chains that may follow upon a superspreading event. Additionally, we demonstrate how a focused stringent testing regime for a subset of “high transmission risk” individuals can effectively control a COVID-19 epidemic for the broader community. Importantly, the extension of our model to heterogenous community dynamics also paves the way for future work that could explicitly model age-structured mixing patterns and infection probabilities by assigning disparate R_0_ values and/or distinct viral load trajectories to different community subgroups. For example, students living in university residence halls may experience a higher daily hazard of infection than older adults in lower density housing (as captured in R_0_), and young adult infections may manifest with lower viral load trajectories that are more likely to present as asymptomatic. Similarly, future modeling efforts could explore variable infection probabilities and/or viral titer trajectories in individuals infected after vaccination or otherwise. Taken together, our model shows the utility of a multi-faceted approach to COVID-19 control and offers a flexible tool to aid in prioritization of interventions in different university or workplace settings.

Finally, our paper presents the only COVID-19 asymptomatic surveillance model published to date that combines asymptomatic testing with contact tracing, thus highlighting the compounding gains effected by these two interventions: contact tracing amplifies the control impacts of both symptom-based and asymptomatic surveillance testing-based isolations, such that even intervention scenarios assuming long delays in isolation after symptom onset or slow turnaround-times for test results can nonetheless greatly reduce the transmission capacity of COVID-19. These findings further emphasize the critical role that asymptomatic surveillance testing will continue to play in ongoing efforts to control COVID-19 epidemics in AY 2021-2022. Even limited asymptomatic surveillance testing can offer substantial gains in case reduction for university and workplace settings with high vaccination rates and/or efficient symptomatic isolation and contact tracing programs in place. Our model allows us to prioritize when and where these gains are most likely to be achieved.

Because we do not explicitly model SARS-CoV-2 transmission in a mechanistic, compartmental framework [69,70], our analysis may overlook some more subtle insights into long-term disease dynamics. More complex analyses of interacting epidemics across larger spatial scales or investigations of the duration of immunity will necessitate implementation of a complete compartmental transmission model. However, our use of a stochastic branching process framework makes our model simple to implement and easily transferrable to other semi-contained small community environments, including a wide range of academic settings and workplaces [26]. We make this tool available to others interested in exploring the impacts of targeted public health interventions—in particular, asymptomatic surveillance testing regimes— on COVID-19 control in more specific settings. We at the University of California, Berkeley are committed to maintaining the safest campus environment possible for our community, using all intervention tools at our disposal. We advise those in similar positions at other institutions to employ the behavioral interventions outlined here, in concert with effective asymptomatic surveillance testing regimes, to reduce community epidemics of COVID-19 in their own communities.

## Data Availability

All code and corresponding data are made available in the article itself and in the cited open-access Github repository.

https://zenodo.org/record/4131223

## Funding

This work was supported by the Miller Institute for Basic Research at the University of California, Berkeley [fellowship to CEB], the Branco Weiss Society in Science at ETH Zurich [fellowship to CEB], a DARPA PREEMPT Cooperative Grant [no. D18AC00031], a COVID-19 Rapid Response Research grant from the Innovative Genomics Institute at the University of California, Berkeley, as well as the NIH [no. R01-GM122061-03] and the NSF EEID program [no. 2011109].

## Supplementary Appendix

### Text S1. *Model Description*

Our publicly-available Github repository (1) provides opensource code to reproduce all simulations and analyses presented in our paper. We summarize the practical implementation details of our modeling design for ease-of-access here.

Our model takes the form of a stochastic branching process model, in which a subset population of exposed individuals (0.5%, derived from the mean percentage of positive tests in our UC Berkeley community (2)) is introduced into a hypothetical 20,000 person community that approximates the campus utilization goals for our university in spring 2021. The model code builds up to a single function ‘replicate.epidemic()’ which runs a specified number of stochastic simulations from a defined parameter set, using the function ‘simulate.epidemic()’. Within the ‘simulate.epidemic()’ function, we first construct a population of 20,000 persons in the sub-function, ‘initiate.pop()’. Within this initiation function, each person in our population is individually numbered, assigned a viral titer trajectory that will be followed if that individual becomes infected (Text B), and assigned a suite of disease metrics drawn stochastically from a specified set of parameter distributions, as outlined in Text S3.

### Text S2. *Within-host viral dynamics*

#### Titer Trajectories

For computational efficiency, we pre-generated 20,000 50-day individual titer trajectories and saved them as an .Rdata file, ‘”titer.dat.20K.Rdata”’. To generate these trajectories, we used a within-host viral kinetics model structured after the classic target cell model (3–5). Code for this model is available in the ‘model-sandbox’ folder of our Github release, under file ‘viral-load.R’, which iterates the following simple model and parameter values derived from Ke et al. (2020), describing the dynamics of SARS-CoV-2 proliferation in the upper respiratory tract (6):

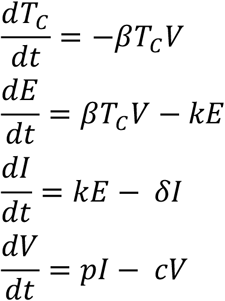

where *T*_*C*_ corresponds to the target cell population, *β* is the transmission rate of free virus to target cell invasion, *k* corresponds to the inverse of the duration of the virus eclipse phase, and *δ* corresponds to the inverse of the incubation period of an infected cell. *p* then gives the burst size of a virus-infected cell and *c* equals the inverse of the lifespan of free virus subject to natural virus mortality and immune predation. Parameter values used to generate each titer trajectory (with a standard deviation of .1x the value of each parameter introduced to add stochasticity in each iteration) are derived from Ke et al. (2020) (6), after fitting this model to individual patient data tracking viral loads through time in the upper respiratory tract of SARS-CoV-2-infected individuals:

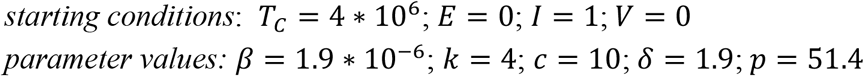

Note that Ke et al. (2020) (6) also explore the within-host dynamics of SARS-CoV-2 infection in the lower respiratory tract; however, since we model human-to-human transmissibility as inferred by viral load in nasopharyngeal swab samples (which better reflect the viral load in the upper respiratory tract), we ignore the lower respiratory dynamics here.

#### Infectivity by Viral Load

After Ke et al. (2020) (6), we next estimated the probability of infection given contact at a specific viral load, using a Michaelis-Menton-like function. Following Ke et al. (2020), we described the probability this probability as:

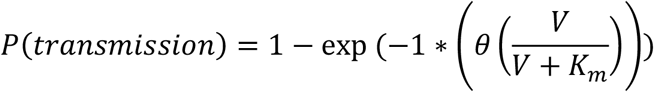

where *K*_*m*_ corresponds to the saturation constant by which proportional gains in infectiousness with viral load diminish at increasingly high viral titers and *θ* is a constant, such that the maximum transmission capacity at any moment equals 1 − *e*^*−θ*^. Ke et al. (2020) modeled a constant hazard of contact events for infectious individuals and therefore fixed *θ* at a value of 0.05, corresponding to a ∼5% probability of a given contact resulting in transmission. Because we draw possible transmissions events from a negative binomial SARS-CoV-2 R_0_ distribution, (mean= 2.5 and *k*=0.10 (7)) but ultimately know that R_E_ for our university environment should have a value of just above one (8), we instead fixed *θ* at a value of 0.72, corresponding to a ∼51% probability of a given contact resulting in transmission, thus effectively halving R_0_ to generate R_E_. The exact probability varied as a function of the timing of each contact event across the trajectory of within-host viral load, with transmissions favored earlier in an infection trajectory when viral load peaks (9).

### Text S3. *Individual disease metrics*

Figures in our paper are derived from 100x replications of each set of parameter values, which we manipulate to explore a range of non-pharmaceutical interventions (NPIs) to combat COVID-19 dynamics in our system. Our flexible model allows for the introduction of NPIs for COVID-19 control in four different forms: (1) group size limits, (2) symptom-based isolations, (3) surveillance testing isolations, and (4) contact tracing isolations that follow after cases are identified through screening from symptomatic or surveillance testing. These interventions modify the suite of disease metrics drawn upon model initiation for each numbered individual in the dataset. We summarize the disease metrics drawn at initiation for all members of the population here:

- **Time of next test:** allocated based on the selected asymptomatic surveillance testing regime. We assume the week starts with day 1 on Saturday and day 7 on Friday. If n.test.days =2, then tests are distributed on Monday (day 3) and Friday (day 7) of each week. As timesteps advance and individuals reach their respective test days, the next test day is updated based on the testing regime (if semi-weekly, the next test day is advanced 3 days; if weekly, the next test day is advanced 7 days; if every-two-weeks, the next test day is advanced 14 days).
- **Beginning/end time of test sensitivity:** based on test limit of detection (LOD) as specified at model outset, this corresponds to the timestep post exposure at which an individual viral titer crosses the threshold for being detectable by the chosen test, both as titers increase at the beginning of a disease trajectory and decrease at the end.
- **Adherence with testing regime:** Y/N, allocated randomly across individuals based on the proportion of the population modeled as complying with the surveillance testing intervention (90% of individuals in all scenarios modeled in our paper).
- **Adherence with group limit**: Y/N, allocated randomly across individuals based on the proportion of the population modeled as complying with the group size limits imposed at outset (90% of individuals in all scenarios modeled in our paper; see ‘number of potential onward cases generated for’ for how group size interacts with cases).
- **Adherence with contact tracing regimen**: Y/N, allocated randomly across individuals based on the proportion of the population modeled as complying with the contact tracing intervention imposed at outset (90% of individuals in all scenarios modeled in our paper).
- **Time of symptom onset**: determined by randomly drawing a titer limit for symptom onset for each individual from a lognormal distribution with a mean of 1e+05 cp/μl RNA and a standard deviation of 1e+04 cp/μl (10–12). The timing of symptom onset then corresponds to the time post-exposure at which each individual’s titer trajectory crosses the corresponding titer limit. According to this approach, under default parameter values, symptom onset occurred between 2 to 4 days post-exposure in our model, and ∼32% of the population never presented with symptoms at all (Fig. 1, main text).
- **Time of symptom-based isolation:** based on delay lag post-symptom onset, drawn from a lognormal distribution with a mean of the specified number of days of symptom isolation lag (1-5 or infinity) and a standard deviation of 0.5 days.
- **Time of tracing-based isolation**: based on contact tracing lag for those adhering to the contact tracing regimen in place. Parameter must be updated with each timestep until individual becomes infected; value then becomes fixed at time of infector isolation, plus corresponding lag drawn from a lognormal distribution with a mean of one day and a standard deviation of 0.5 days.
- **Time of testing-based isolation:** based on turnaround time to isolation post testing, drawn from a lognormal distribution with a mean of the specified number of delay days (1-5, 10, or infinity) and a standard deviation of 0.5 days. Parameter is updated when ‘time of next test’ is updated for each individual in our model.
- **Disease status:** ‘susceptible’ = 0, ‘exposed’ = 3, ‘infectious’ = 1, ‘recovered’ = 5, ‘vaccinated’= 6. At onset, all individuals are modeled as susceptible, excepting the 0.5% which are introduced as infectious (1) to seed the epidemic and the ‘prop-vaccinated’, a parameter encoding the proportion of the target population that is vaccinated prior to the start of epidemic simulations. We additionally encode a ‘prop-breakthrough’ parameter which corresponds to the proportion of vaccinated individuals who experience breakthrough infections. In simulations presented in our paper, 95% of vaccinated individuals are treated as if fully immune, while 5% of individuals experience breakthrough infections; these breakthrough cases are modeled stochastically, based on probability at the timestep in which each possible infection encounter occurs.

#### Number of potential onward cases generated

Several figures in the main text of our manuscript present the R_E_ reduction capacity of a specified intervention, which we calculate as the difference between the average of the number of potential onward cases generated and the number of actual onward cases generated for each individual after an intervention is adopted. To compute the number of potential onward cases generated for each individual, we first draw a number of possible cases from a negative binomial distribution with a mean of 2.5 and a dispersion parameter (k) of 0.10, as estimated for SARS-CoV-2 (7) (or with a mean of 6 in later simulations to represent the heightened transmissibility of the Delta variant (13)). Next, we assume that a minority of transmission events will be lost to the external environment through contacts between UC Berkeley students and members from the outside community. We do not track these ‘lost cases’ but instead simply reduce the total number of potential onward cases to the proportion constrained within UCB: 90% in simulations presented in the main text and 50% in the sensitivity analysis presented in Fig. S5.

Then, we draw a number of possible onward transmission events for the remaining cases for each infectious individual from a simple Poisson distribution with *λ* = 3, signifying the average number of possible encounters (i.e. cross-household dining, shared car rides, indoor meetings, etc.) per person that could result in transmission. We then distribute each infectious person’s original number of R_0_-derived potential cases among these events at random, ensuring that multiple transmissions are possible at a single event; the most extreme superspreading events thus occur when persons with heterogeneously high infectiousness draw a large number of potential cases, which are concentrated within a relatively small number of discrete transmission events. For example, if an infectious individual draws an R_0_ value of 16 and an event number value of 4, then those 16 potential infections are randomly distributed among 4 events.

Next, we use published estimates of the generation time of onward transmission events for SARS-CoV-2 infection to draw event times for each event, based on a weibull distribution with a shape parameter = 2.826 and a scale parameter = 5.665, as specified in Ferretti et al. (2020) (9). Following the above example, 4 discrete generation times would be assigned to cases across the 4 pre-allocated events.

Since each individual is already pre-assigned a within-host viral titer trajectory in our modeling framework, we next examine the viral load specified at the generation time of each transmission event and determine if each case assigned to that event actually occurs. Each case is considered individually, and the probability of transmission is computed stochastically based on the value of the individual’s viral titer at the time of the event (higher titer infections are more likely to generate onward transmission events) (Text S2). In the above example, all 16 possible transmissions would be individually assessed, though several would have the same titer, corresponding to the infectious person’s titer at the time point of each contact event (4 possible). Since our maximum probability of a case occurring at max viral load is ∼51% (Text S2), our original R_0_-derived cases are here halved, resulting in an average of 1.05 onward transmission events per infectious individual (or just under 3 in the case of Delta simulations) in the absence of the NPIs examined here (but reflecting social distancing and mask wearing), which, as specified in the main text, is in line with current estimates from Alameda County, CA (8).

For the purposes of our example, let’s assume that 10 of those possible 16 cases occur, allocated across 4 different events, with 7 cases at one event and one case each at 3 other events.

- **Number of actual onward cases generated**: From the number of possible cases generated, we next apply the relevant intervention and iterate forward in time to determine the actual number of cases generated by each infectious individual across the time course of our modeled epidemics. For symptom and surveillance testing-based isolations, as well as contact tracing, no cases are generated if an infectious individual is isolated prior to the generation time of any possible onward cases. For NPIs in the form of group size limits, case reduction in our model is performed prior to the initiation of the epidemic time series, and case numbers for each transmission event are truncated at the intervention limit.

Again following the example listed above, if we imagine that the imposed group size limit is 6, then the 7 cases assigned to a single event will be truncated to 6, meaning that 9 out of the 10 potential cases are allowed to occur after the intervention. Our model is conservative in assessing the impact of a group-size intervention by allowing some portion of those superspreading cases to occur, rather than assuming that a group size limit-abiding infectious individual does not attend larger-than-allowable events altogether. Because only 90% of the population adheres to group size intervention in any given simulation (or 50% in sensitivity analyses; see Fig. S1), some proportion of large superspreading events will still take place at random, even after NPIs are imposed.

Following onset of infection, the timings of symptom-, tracing-, and asymptomatic testing-based isolations are then compared and the earliest time is selected as the actual mechanism (if any) of isolation for that individual. The number of actual onward cases generated is then updated if isolation occurs prior to some new case generations. Additionally, all individuals identified as infectious are additionally assigned the following metrics:

- **Isolation time of infector**
- **Source of infection** (external Alameda County vs. UC Berkeley community member)
- **ID number of infector**, if from UC Berkeley

The cycle then repeats in the next timestep when all “actual infections” for each infectious individual are then assigned to new susceptible individuals. The epidemic continues with updated parameters for all newly exposed individuals until either the end of the time series is reached or no more susceptible individuals remain in the population.

## Supplementary Figures

**Figure S1.**
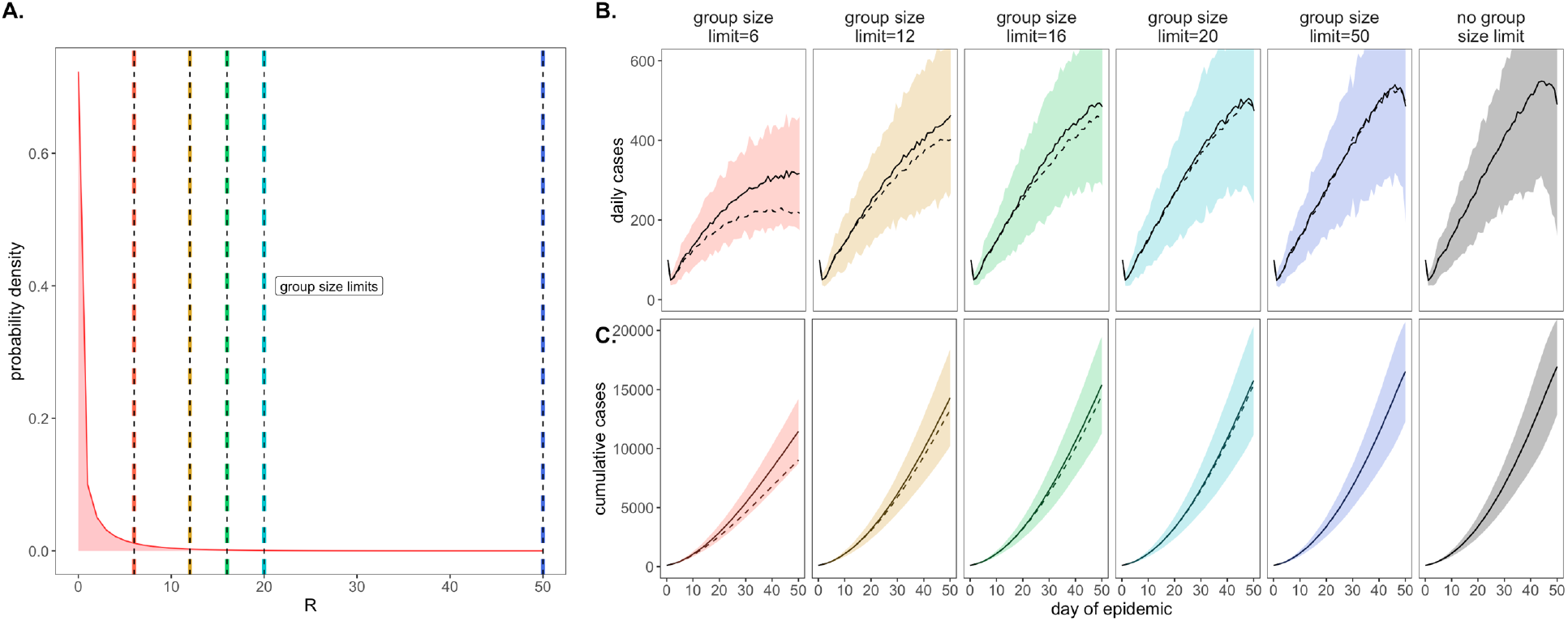
Figure replicates Fig. 2 (main text) assuming only 50% adherence to group size limitations vs. the 90% adherence presented in the main text. **A**. Negative binomial R_E_ distribution with mean = 1.05 and dispersion parameter (k) = 0.10. The colored vertical dashes indicate group size limits that ‘chop the tail’ on the R_E_ distribution; for 90% of the population, coincident cases allocated to the same transmission event were truncated at the corresponding threshold for each intervention. **B**. Daily new cases and, **C**. Cumulative cases, across a 50-day time series with 95% confidence intervals by standard error depicted under corresponding, color-coded group size limits. Mean output of simulations under 50% adherence are shown as solid black lines, with the dashed line corresponding to mean output under the 90% adherence assumptions presented in the main text.

**Figure S2.**
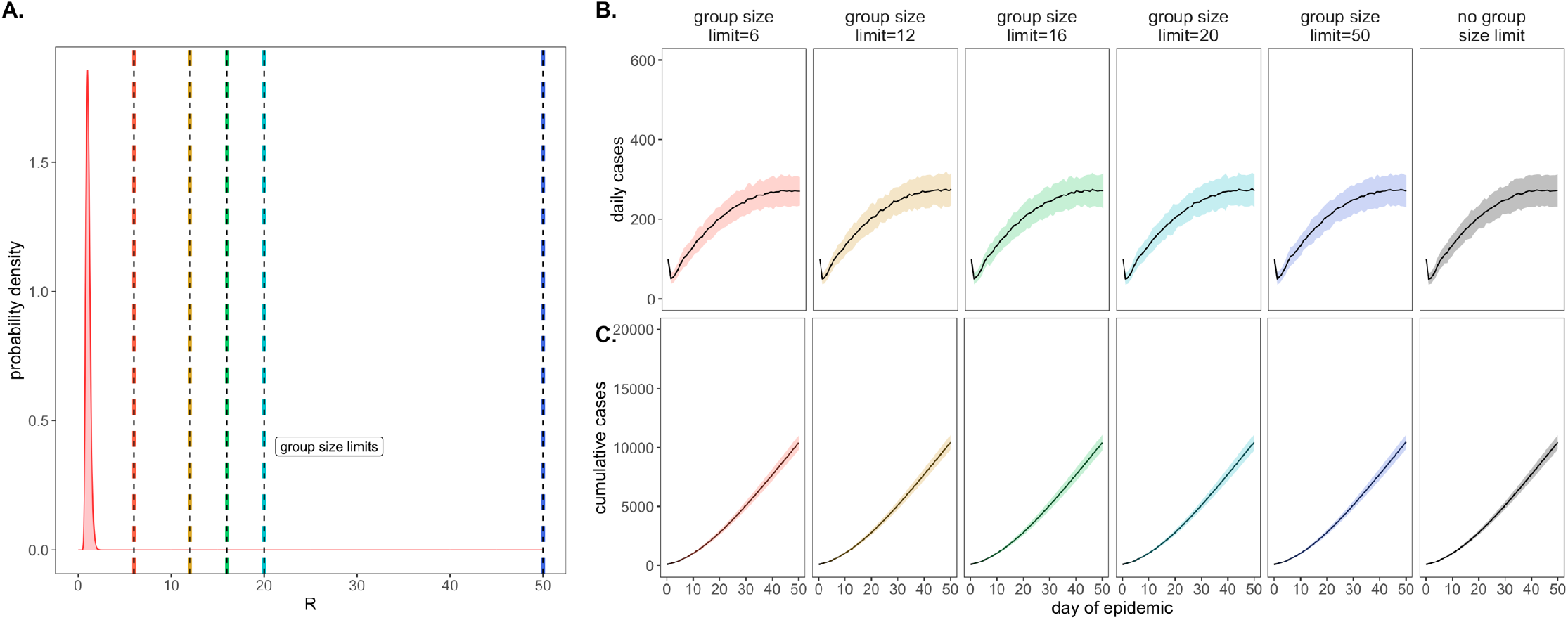
Figure replicates Fig. 2 (main text) at a log-normal distribution for R_E_, instead of negative binomial. **A**. Log-normal R_E_ distribution with a mean of 1.05 and a standard deviation of 1.233. The colored vertical dashes indicate the group size limits that ‘chop the tail’ on the R_E_ distribution. **B**. Daily new cases and, **C**. cumulative cases, across a 50-day time series under corresponding, color-coded group size limits.

**Figure S3.**
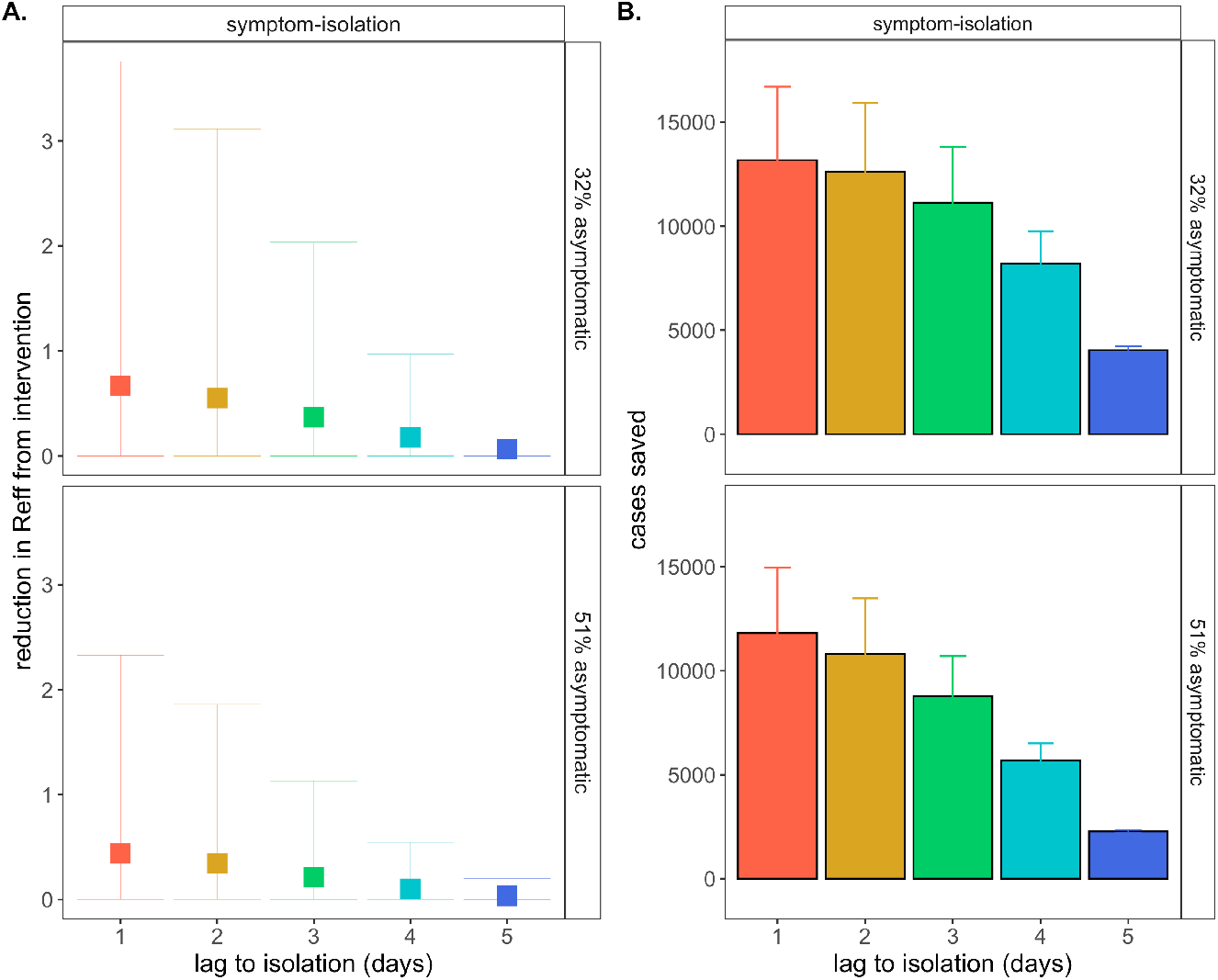
Figure replicates symptom-isolation panels from Fig. 3 (main text) in top row, showing **A**. mean reduction in R_E_ and **B**. cumulative cases saved across 50-day simulated epidemics under differing lag times to isolation, assuming a threshold titer for symptom onset by which ∼32% of the population presents as asymptomatic. A comparison at a titer threshold for which ∼51% of the population presents as asymptomatic demonstrates how a higher proportion of asymptomatic individuals in the population erodes the effectiveness of the symptom-based isolation intervention; asymptomatic status has no impact on the effectiveness of group size limits or asymptomatic surveillance testing interventions.

**Figure S4.**
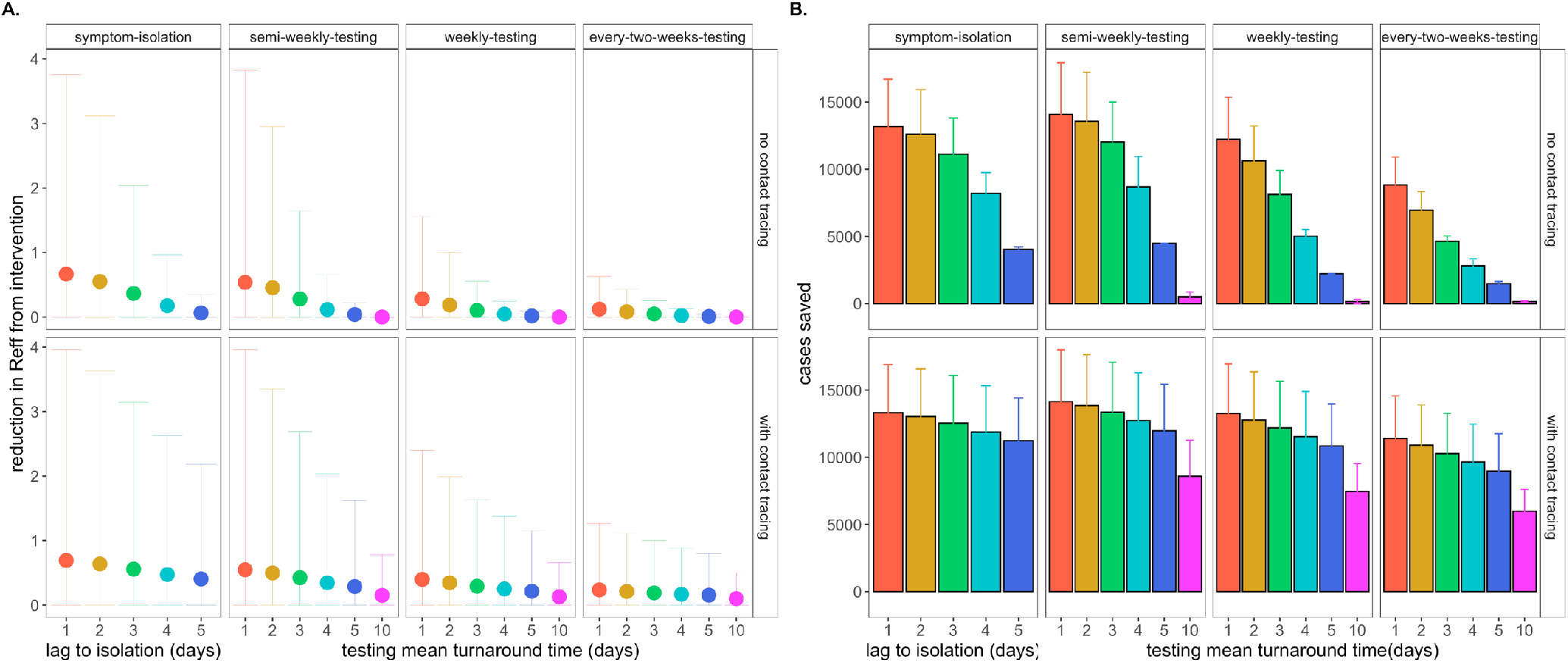
Figure replicates symptom-isolation panels from Fig. 3 (main text) in top row, showing **A**. mean reduction in R_E_ and **B**. cumulative cases saved across 50-day simulated epidemics for NPIs of both symptom-based and testing-based isolation, across a range of different lag times or turnaround times to isolation (for, respectively symptom-or testing-based isolations). All testing-based interventions depicted are shown at a limit of detection=10^1^ cp/μl. In the bottom row, **A**. mean reduction in R_E_ and **B**. cumulative cases saved are depicted for a comparative intervention which adds an additional single-day lag in contact tracing to the respective symptom-based or testing-based isolation. Under these combined interventions, even previously ineffective testing interventions with 10-day turnaround time show gains beyond no intervention at all.

**Figure S5.**
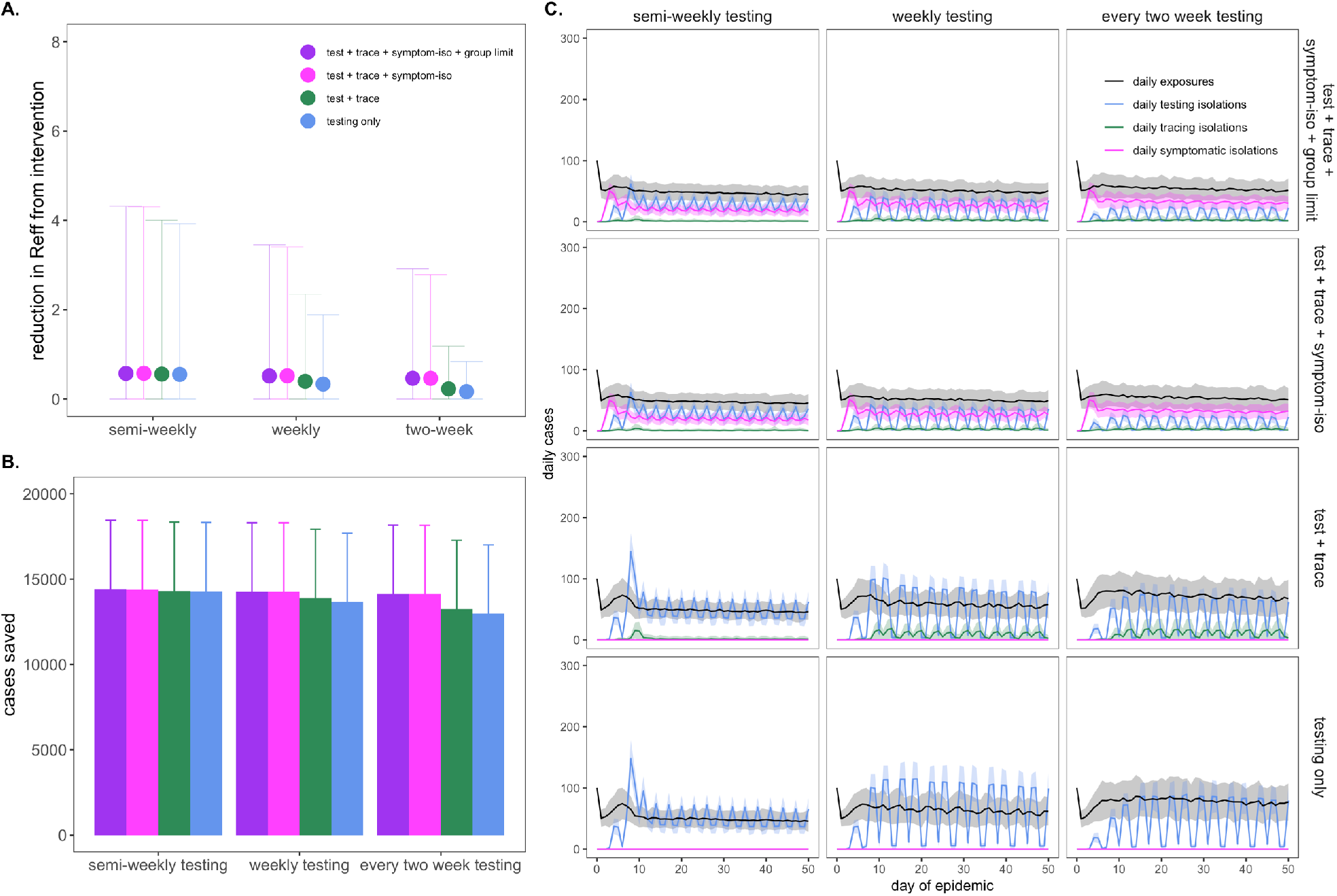
Figure replicates Fig. 4 of the main text, under assumptions of 50% of cases lost to the outside community, as compared to the 10% modeled in the main article. **A**. Mean reduction in R_E_, **B**. cumulative cases saved, and **C**. daily case counts for the first 50 days of the epidemic, across regimes of differing testing frequency and a combination of surveillance testing, contact tracing, symptomatic isolation, and group size limit interventions. All scenarios depicted here assumed test turnaround time, symptomatic isolation lags, and contact tracing lags drawn from a log-normal distribution with mean=one day. Limit of detection was fixed at 10^1^ and group size limits at 12. Dynamics shown here are from simulations in which testing was limited to two test days per week. NPIs have proportionally less impact on R_E_ reduction (A) but nonetheless manage to avert an equal number of cases (B) when the university is modeled as a more open, community-integrated environment. Under this scenario, interventions function primarily to isolate cases from the external environment, rather than curb onward, within-community transmission. For this reason, daily variance in exposure rate is also diminished under assumptions of a higher proportion of transmissions lost to the surrounding community. *Note: R_E_ reduction (panel A) is calculated as the difference in mean R_E_ in the absence vs. presence of a given NPI. The upper confidence limit (uci) in R_E_ reduction is calculated as the difference in uci R_E_ in the absence vs. presence of NPI. In our model, mean R_E_ in the absence of NPI equals 1.05 and uci R_E_ in the absence of NPI equals 8.6.

**Figure S6.**
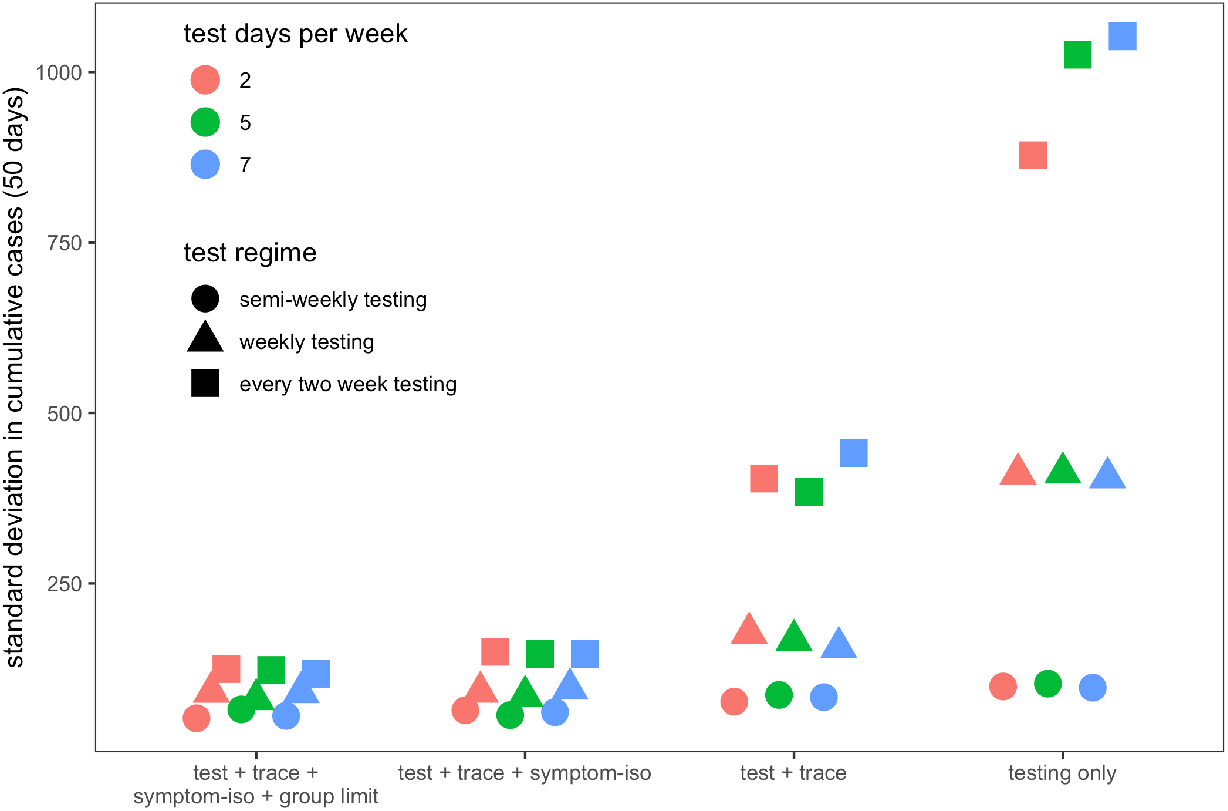
Figure extends results from Fig. 4 (main text), showing the standard deviation in cumulative cases from 50-day simulated epidemics, across regimes of differing testing frequency and a combination of surveillance testing, contact tracing, symptomatic isolation, and group size limit interventions. All scenarios depicted here assume test turnaround time, symptomatic isolation lags, and contact tracing lags drawn from a log-normal distribution with mean=1 day. Limit of detection is fixed at 10^1^ and group size limits at 12. Dynamics compare tests of differing frequency (semi-weekly, weekly, every two weeks) distributed across variable numbers of days in a given week (2,5,7). Additional layers of intervention and more testing days per week reduce the standard deviation in cumulative cases.

**Figure S7.**
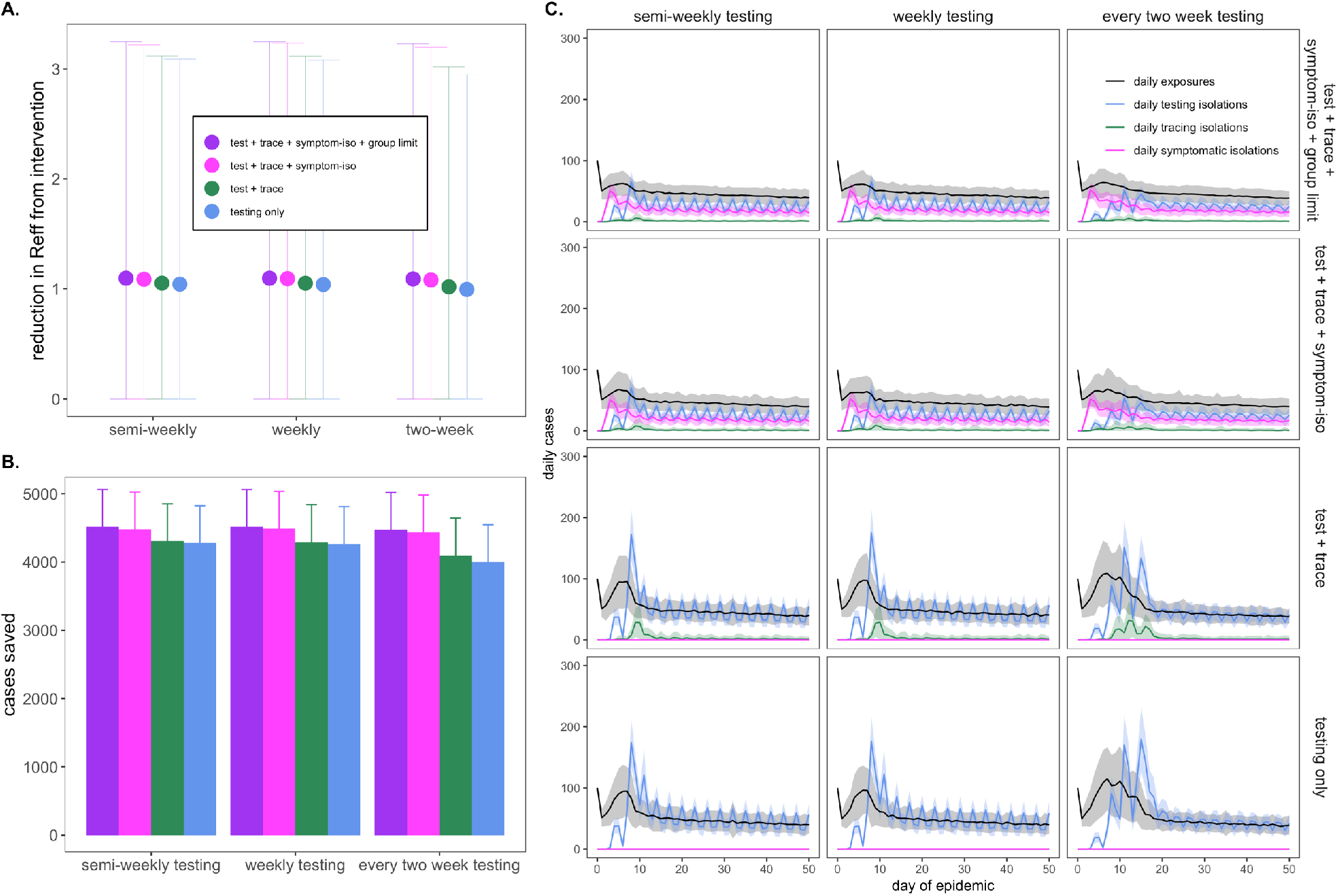
Figure largely replicates Fig. 4 of the main text, under assumptions of mean R_0_ = 6 and 60% of the baseline campus population vaccinated, approximating circulation of the Delta variant in the undergraduate population of the University of Alabama, Tuscaloosa at the time of this writing. Note that y-axes for panel **A**. and **B**. differ from those depicted in Fig 4 of the main text and from Fig. S8 below. **A**. Mean reduction in R_E_ and **B**. cumulative cases saved compared to a baseline scenario in which no behavior-based or testing NPIs were applied but simulations were run under assumptions of 60% vaccination in an R_0_=6 environment. **C**. Daily case counts for the first 50 days of the epidemic, across regimes of differing testing frequency and a combination of surveillance testing, contact tracing, symptomatic isolation, and group size limit interventions. All scenarios depicted here assumed test turnaround time, symptomatic isolation lags, and contact tracing lags drawn from a log-normal distribution with mean=one day. Limit of detection was fixed at 10^1^ and group size limits at 12. Dynamics shown here are from simulations in which testing was limited to two test days per week. Combined, asymptomatic surveillance testing and behavior-based NPIs still reduce R_E_ and avert cases but impacts are reduced compared to no vaccination settings (main text) because fewer opportunities for infection arise. Variance between simulations and interventions is also diminished in this more mild epidemic scenario, indicating that testing alone, without rigorous extensive additional interventions, can effectively control outbreaks. *Note: R_E_ reduction (panel A) is calculated as the difference in mean R_E_ in the absence vs. presence of a given NPI. The upper confidence limit (uci) in R_E_ reduction is calculated as the difference in uci R_E_ in the absence vs. presence of NPI. In our model, mean R_E_ under Delta variant transmission assumptions in the absence of NPIs, but including 60% population-level vaccination, equals 1.12 and uci R_E_ equals 3.33.

**Figure S8.**
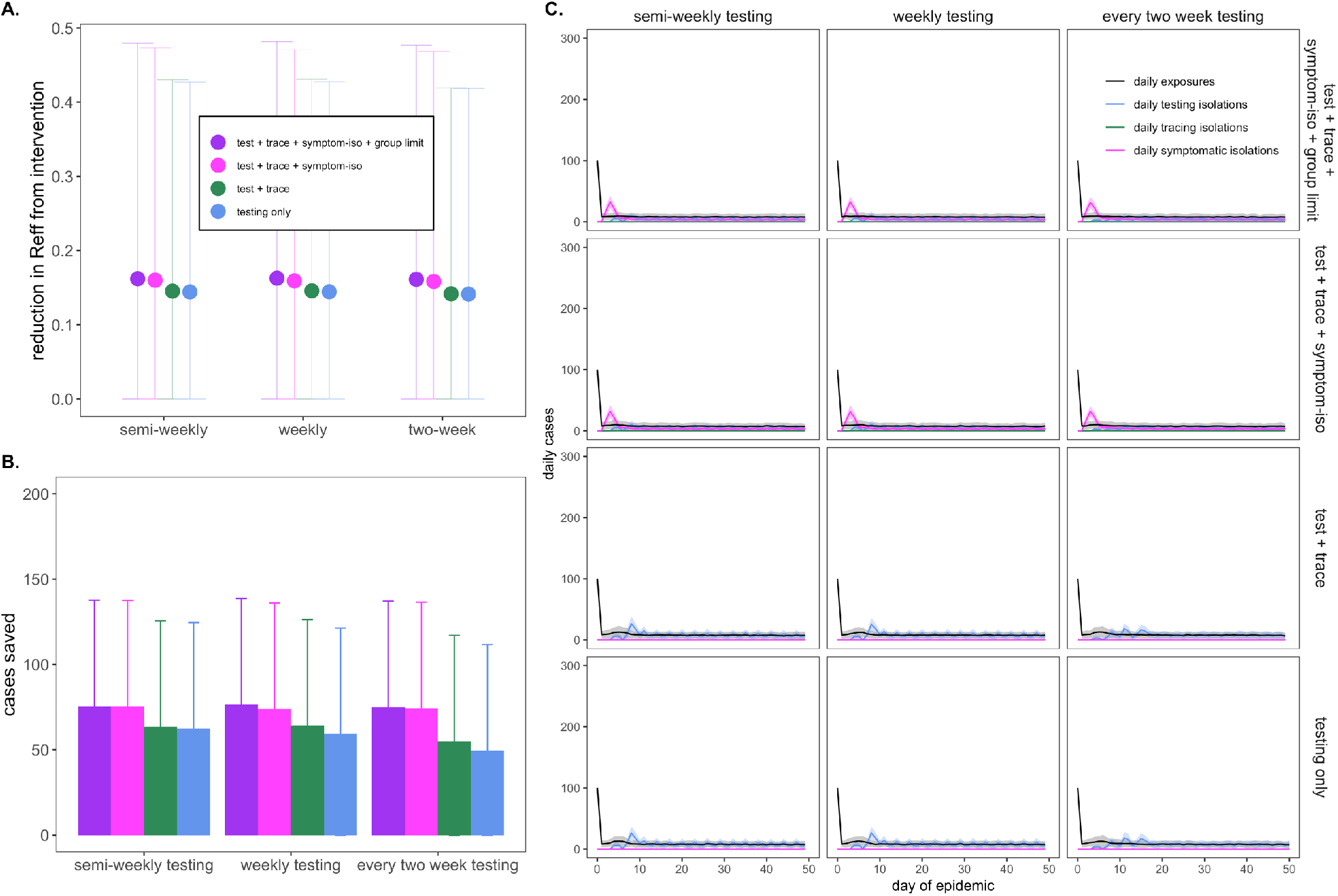
Figure largely replicates Fig. 4 of the main text, under assumptions of mean R_0_ = 6 and 97.7% of the baseline campus population vaccinated, approximating circulation of the Delta variant in the undergraduate population of UC Berkeley at the time of this writing. Note that y-axes for panel **A**. and **B**. differ from those depicted in Fig 4 of the main text and from Fig. S7 above. **A**. Mean reduction in R_E_ and **B**. cumulative cases saved compared to a baseline scenario in which no behavior-based or testing NPIs were applied but simulations were run under assumptions of 97.7% vaccination in an R_0_=6 environment. **C**. Daily case counts for the first 50 days of the epidemic, across regimes of differing testing frequency and a combination of surveillance testing, contact tracing, symptomatic isolation, and group size limit interventions. All scenarios depicted here assumed test turnaround time, symptomatic isolation lags, and contact tracing lags drawn from a log-normal distribution with mean=one day. Limit of detection was fixed at 10^1^ and group size limits at 12. Dynamics shown here are from simulations in which testing was limited to two test days per week. Even in highly vaccinated university settings, behavior-based NPIs and asymptomatic surveillance testing reduce R_E_ and avert cases largely derived from breakthrough infections, though lower baseline case counts equate to lower gains in R_E_ reduction and case aversions. Variance between simulations and between interventions is most diminished in this epidemic scenario, indicating that testing alone, without rigorous extensive additional interventions, can effectively control outbreaks. *Note: R_E_ reduction (panel A) is calculated as the difference in mean R_E_ in the absence vs. presence of a given NPI. The upper confidence limit (uci) in R_E_ reduction is calculated as the difference in uci R_E_ in the absence vs. presence of NPI. In our model, mean R_E_ under Delta variant transmission assumptions in the absence of NPIs, but including 97.7% population-level vaccination, equals 0.17 and uci R_E_ equals 0.51.

## Legends for Datasets S1-S3

**Dataset S1. Averaged total cases saved and mean R**_**E**_ **reduction across group size limit, symptomatic isolation, and surveillance testing NPIs**. Summarized model output from 100x simulations across all NPIs presented in Fig. 2 and Fig. 3, main text. Confidence intervals represent 1.96*standard deviation in case reduction or R_E_ reduction.

**Dataset S2. Averaged total cases saved and mean R**_**E**_ **reduction across symptomatic isolation, and surveillance testing NPIs, under regimes with and without contact tracing**. Summarized model output from 100x simulations across all NPIs presented in SI-Appendix, Fig. S3.

**Dataset S3. Averaged total cases saved and mean R**_**E**_ **reduction across combined intervention approaches**. Summarized model output from 100x simulations across all NPIs presented in Fig. 4, main text.

All other model output available as saved .Rdata files in our publicly-available Github repository:

Brook CE, Northrup GR, Boots M (2020) Code for “Optimizing COVID-19 control with asymptomatic surveillance testing in a university environment.” doi:10.5281/zenodo.4131223

## Notes

### Competing Interest Statement

The authors have declared no competing interest.

### Funding Statement

CEB was funded by the Miller Institute for Basic Research at the University of California, Berkeley, the Branco Weiss Society in Science Fellowship from ETH Zurich, a DARPA PREEMPT Cooperative Grant (no. D18AC00031), and a COVID-19 Rapid Response Research grant from the Innovative Genomics Institute at the University of California, Berkeley. MB was supported by NIH grant no. R01-GM122061-03 and NSF EEID grant no. 2011109.

### Author Declarations

No IRB approval was necessary for this study.

### Summary of Updates

Sensitivity analyses requested by reviewers at Epidemics were added (Fig S1, S5, S7, S8), as was Table 1, and scenarios run incorporating Delta transmission and partially vaccinated university settings.

